# Maternal *APOL1* Genotypes and Preeclampsia Risk

**DOI:** 10.64898/2026.03.30.26349770

**Authors:** Wen Tong, Frances Conti-Ramsden, Hannah Beckwith, Argyro Syngelaki, Ioannis Mitrogiannis, Lucy Chappell, Pirro Hysi, Catherine Williamson, Sophie Limou, Kypros Nicolaides, Kate Bramham, Antonio de Marvao

## Abstract

**Background:** APOL1 risk alleles are prevalent in individuals of West African ancestry and associated with increased risk of kidney disease. Although preeclampsia disproportionately affects women of Black ethnic backgrounds, evidence linking APOL1 alleles to preeclampsia remains conflicting.

**Objectives:** The purpose of this study was to explore whether maternal APOL1 alleles contribute to preeclampsia risk and associated adverse pregnancy outcomes.

**Study design:** We conducted a nested case-control study of 5210 pregnant women, including 745 preeclampsia cases and 949 controls of Black self-reported ethnicity, 1385 preeclampsia cases and 2131 controls of White self-reported ethnicity. APOL1 G1 and G2 risk alleles were directly genotyped on the Illumina Infinium® Global Screening Array. Associations with preeclampsia, early preeclampsia, recurrent preeclampsia, birthweight centiles and gestational age at delivery were examined using regression models assuming a recessive mode of inheritance with adjustment for established risk factors and stratification by self-reported ethnicity and genetically-determined ancestry.

**Results:** Presence of APOL1 risk alleles was almost exclusively observed in women of Black self-reported ethnicity. 168/949 controls (17.7%) and 133/745 cases (17.9%) carried two APOL1 risk alleles, and these women did not have a significantly increased risk of preeclampsia compared to those with zero or one APOL1 risk alleles in adjusted analyses (OR 1.00, 95% CI 0.76-1.29, p=0.972). When restricting analysis to women of Black self-reported ethnicity only, no association was observed between APOL1 genotype and preeclampsia risk (adjusted OR 0.94, 95% CI 0.61-1.25, p=0.673). When restricting analysis to women of pan-African genetically-determined ancestry only, also no association was observed between APOL1 genotype and preeclampsia risk (adjusted OR 1.00, 95% CI 0.76-1.32). No associations were found between number of APOL1 risk alleles and early preeclampsia, recurrent preeclampsia, birthweight centile or gestational age at delivery after adjustment for established risk factors and stratification by self-reported ethnicity or genetically-determined ancestry.

**Conclusions:** Maternal APOL1 risk alleles do not independently influence preeclampsia risk or related adverse outcomes in a multi-ethnic pregnancy study. Future studies should examine whether fetal APOL1 genotypes, alone or in interaction with maternal genotypes, contribute to preeclampsia risk.

**At a Glance:** *Why was this study conducted?:*

- APOL1 risk alleles are prevalent in West African ancestry populations and associated with kidney disease
- Preeclampsia disproportionately affects women of Black ethnicity
- Previous studies showed conflicting evidence linking maternal *APOL1* alleles to preeclampsia risk

*What are the key findings?:*

- In 5,210 multi-ethnic women, maternal *APOL1* risk alleles showed no independent association with preeclampsia or related adverse outcomes
- Associations in unadjusted analyses reflected West African genetic ancestry rather than direct causation

*What does this study add to what is already known?:*

- Largest genetic study of maternal *APOL1* and preeclampsia in women of Black ethnicity (745 cases)
- Demonstrates *APOL1* alleles serve as ancestry markers rather than causal variants
- Highlights need to examine fetal genotypes and gene-environment interactions

## Introduction

Preeclampsia affects 2-8% of pregnancies worldwide and remains a leading cause of maternal and perinatal mortality and morbidity, with 76,000 maternal and 500,000 perinatal deaths every year[1,2]. Women of Black ethnic backgrounds are at increased risk of developing preeclampsia with consistently worse outcomes reported in comparison to women of White ethnic backgrounds[3-10]. Established cardiovascular risk factors and sociodemographic deprivation contribute to, but do not fully explain these disparities[11,12].

There is growing interest in the role of *APOL1* risk alleles in preeclampsia in individuals of recent West African ancestry. *APOL1* alleles G1 and G2 confer protection against African sleeping sickness transmitted by *Trypanosoma brucei rhodesiense* when compared with the reference G0 ancestral allele[13,14]. These alleles are almost exclusive to individuals of recent West African ancestry, with >50% of African Americans carrying one or two risk alleles[13,15]. *APOL1* risk genotypes are strongly associated with non-diabetic kidney disease and account for much of the excess burden of kidney disease in people of recent West African descent[13,15-19]. Given the well-established bidirectional relationship between kidney disease and preeclampsia, *APOL1* has emerged as a candidate contributor to preeclampsia risk.

Whilst *in vitro* and animal studies have suggested a role of *APOL1* in preeclampsia pathogenesis[20-22], human evidence on maternal *APOL1* risk alleles and preeclampsia remains inconclusive[23]. The largest human studies to date have included fewer than 500 women of Black ethnic backgrounds, highlighting the need for larger, well-powered studies[23-26].

The aim of this study was to examine the association between maternal *APOL1* risk alleles and risk of preeclampsia, early preeclampsia, recurrent preeclampsia and preeclampsia-related outcomes in a large, contemporaneous, multi-ethnic study.

## Materials and Methods

### Study Design

The Harris Birthright Cohort is a prospective observational cohort study recruiting women with singleton pregnancies who attend for routine hospital visits at King’s College Hospital, London, and Medway Maritime Hospital, Kent (REC: 02-03-033, IRAS:89351). Participants of this sub-study were enrolled 2006-2021. Following informed consent, participants had longitudinal assessments at 12- and 20 weeks’ gestation with ultrasound measurements and blood sampling. Maternal ethnicity was self-reported from multiple choice options, and only participants who self-identified as either of Black or White were included. Pregnancy outcomes were curated by clinicians through detailed review of patient records, laboratory investigations and ultrasound data. Preeclampsia was defined according to the 2019 American College of Obstetricians and Gynecologists criteria[27]. Women with preeclampsia were matched to controls, with matching by self-reported ethnicity (SRE) and date of recruitment.

Early, preterm and term preeclampsia were defined as preeclampsia with delivery before 34, between 34 and 37, and after 37 weeks gestational age, respectively. Recurrent preeclampsia was defined as preeclampsia with previous history of preeclampsia, and parous women without a history of preeclampsia were defined as the control group.

### Sample Processing and Genotyping

Blood samples were centrifuged and buffy coat layers were collected and preserved at -80 °C until analysis. Following genomic DNA extraction, samples underwent quantification, normalization and genotyping using the Illumina Infinium® Global Screening Array version 3.0 with multi-disease add-on (∼ 700,000 variants, comprehensive genome-wide coverage optimized for diverse ethnic populations). *APOL1* G1 (rs73885319, p.S342G, and rs60910145, p.I384M) and G2 (rs71785313, p.N388_Y389del) renal risk alleles were directly genotyped [28]. Other *APOL1* non-risk genotypes were collectively assigned the G0 genotype.

Genotype clustering was performed using Illumina’s GenCall algorithm within Genome Studio 2.0 software, with sample quality control measures applied following established GWAS protocols[29]. Quality control procedures included screening for undisclosed familial relationships and verification of concordance between self-reported and genetically-determined sex. All quality control steps were performed in PLINK 1.9 (www.cog-genomics.org/plink/1.9/)[30].

### Genetic Ancestry Determination

Individual genetic ancestry was determined as previously described through supervised ancestry analysis using the ADMIXTURE software[31-33]. Ancestry estimates derived from reference populations were consolidated into five continental superpopulations: Pan-African (AFR), Admixed American (AMR), East Asian (EAS), European (EUR), and South Asian (SAS). Predominant genetically-estimated ancestry group was assigned as follows; >50% pan-African genetic ancestry – AFR, >50% European genetic ancestry – EUR. Admixed ancestry was assigned if neither AFR nor EUR genetic ancestry reached 50% in total. Congruence between predominant genetically-determined ancestry (GDA) and SRE was assessed: women with Black SRE and >50% AFR genetic ancestry, and women with White SRE and >50% EUR genetic ancestry were considered concordant[34].

### Statistical Analysis

The following *APOL1* genotypes were assigned from the individual genotype data: G1G1, G2G2, G1G2, G1G0, G2G0 or G0G0. Analysis was primarily performed using a recessive model of inheritance of *APOL1* risk alleles, in which odds of the outcome in the presence of two risk alleles (G1G1, G1G2 or G2G2) were compared to zero or one risk alleles (G1G0, G2G0 or G0G0). Secondary analyses assuming additive (presence of two AND one risk allele compared to zero risk alleles) and dominant (presence of two OR one risk alleles compared to zero risk alleles) models of inheritance of *APOL1* risk alleles are reported in the Supplementary Material[15]. The association between *APOL1* risk alleles and the primary outcomes (preeclampsia, early preeclampsia and recurrent preeclampsia) and secondary outcomes (gestational age at delivery and birthweight centile) was assessed using logistic and linear regression models, as appropriate. For secondary outcomes, regression analyses were stratified by case control status.

To assess whether there was an association between *APOL1* risk alleles and primary and secondary outcomes independent of established risk factors for preeclampsia, we used the following models: i) unadjusted model, ii) model adjusted for maternal age and body mass index (BMI) and iii) clinical risk factor model with adjustment for the following maternal risk factors: maternal age, BMI, conception method (spontaneous, ovulation induction or *in vitro* fertilization (IVF)), smoking status, chronic hypertension, diabetes mellitus (Type 1 or Type 2), maternal history of preeclampsia, nulliparity and previous preeclampsia[35].

For linear regression models, nested model comparisons were performed using F-tests via ANOVA to determine whether the addition of covariates significantly improved model fit compared to the unadjusted model. For logistic regression models, nested models were compared using likelihood ratio tests, evaluated against the chi-square distribution. Akaike Information Criterion (AIC) values were calculated for all models, with lower AIC indicating better model fit whilst accounting for model complexity.

All data cleaning and analysis was carried out in R version 4.4.3 (R Foundation for Statistical Computing, Vienna, Austria)[36].

## Results

### Study Characteristics and *APOL1* Risk Allele Distribution

Of 5,675 genotyped individuals, 5,210 passed genomic quality control steps and had complete covariate and outcome data (Figure S1). Maternal baseline characteristics, risk factors for preeclampsia and pregnancy outcomes stratified by SRE and GDA are shown in Tables 1 and S1, respectively. There was a higher proportion of women with preeclampsia of Black SRE (n=745, 44.0%) compared to women of White SRE (n=1385, 39.4%). Early and recurrent preeclampsia were more common in women of Black SRE (n=115, 15.4% and n=116, 15.6%, respectively) compared to women of White SRE (n=124, 9.0% and n=147, 10.6%, respectively).

**Table 1.**
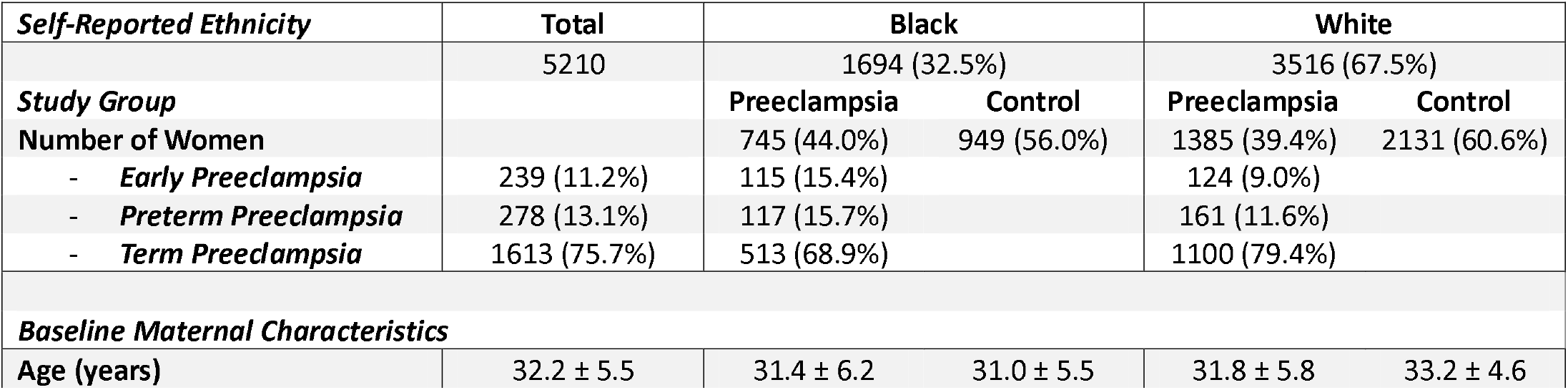
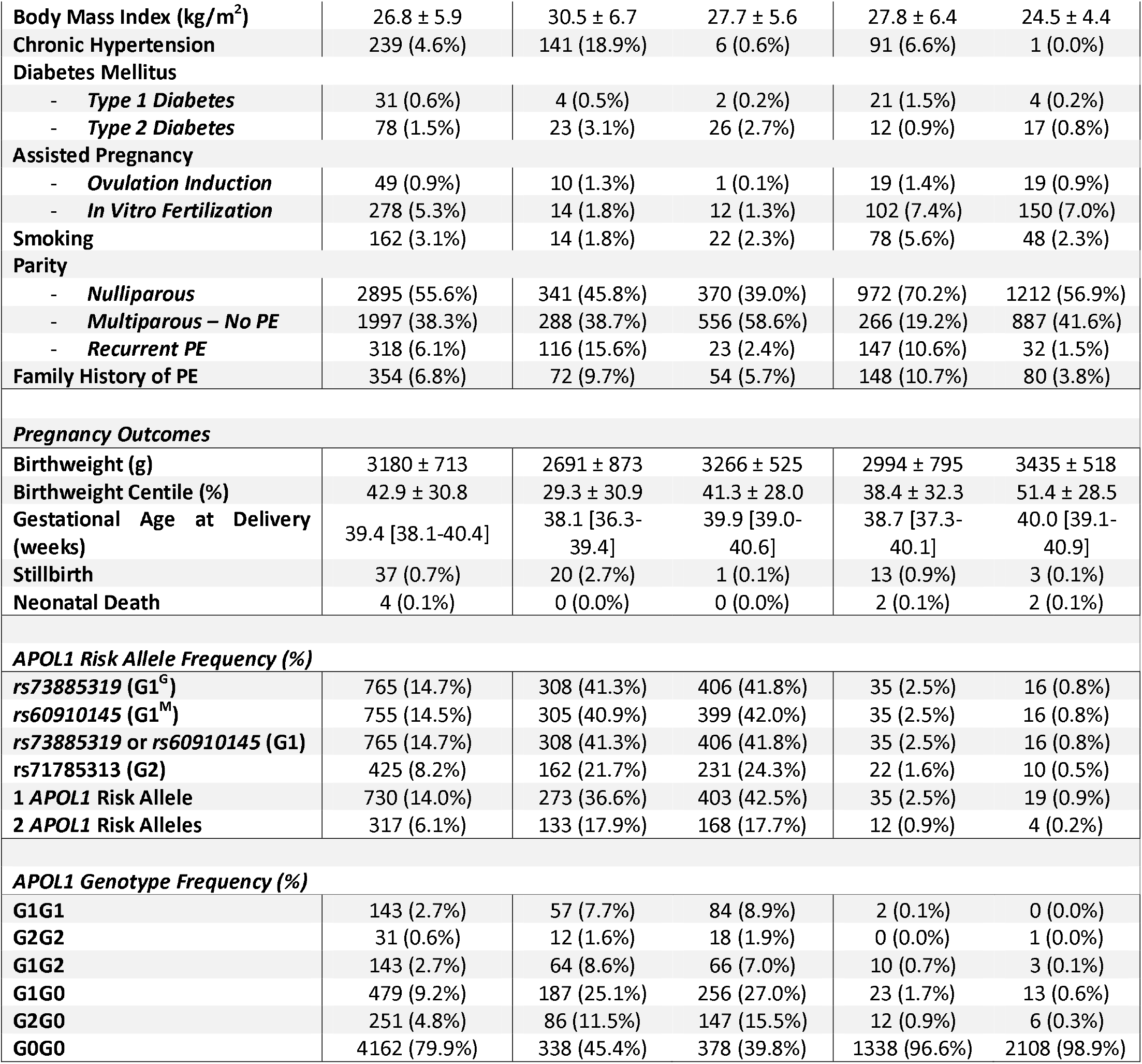
Baseline Characteristics, Clinical Risk Factors for Preeclampsia, *APOL1* Risk Allele Distribution in Women with Preeclampsia and without Preeclampsia Stratified by Self-Reported Ethnicity. Results are presented as mean ± standard deviation or median [interquartile range] for continuous variables and count (percentage) for categorical variables. Abbreviations: PE = preeclampsia; Definitions: Early preeclampsia = delivery with preeclampsia before 34 weeks gestational age, preterm preeclampsia = delivery with preeclampsia between 34 and 37 weeks gestational age, term preeclampsia = delivery with preeclampsia after 37 weeks gestational age.

Distribution of *APOL1* allele frequencies and genotypes by SRE and study group are shown in Table 1. rs73885319 (G1^G^) and rs60910145 (G1^M^) risk alleles showed near perfect (r^2^=0.987) linkage disequilibrium. Occurrence of two *APOL1* risk alleles was common in women of Black SRE (n=168, 17.7% and n=133, 17.9% in controls and preeclampsia, respectively), but rare in women of White SRE (n=4, 0.2% and n=12, 0.9% in controls and preeclampsia, respectively).

GDA distribution in women of Black and White SRE is shown in Figures 1A and 1B, respectively. Congruence between SRE and GDA was 92.8% across the entire study. Notably, of the 16 women of White SRE with two *APOL1* risk alleles, only one woman had >50% EUR genetic ancestry, whilst 15 had >50% AFR genetic ancestry. Women with >50% AFR genetic ancestry had an average of 92.6% pan-African genetic ancestry overall, 70.0% West African genetic ancestry and 8.3% East African genetic ancestry.

**Figure 1.**
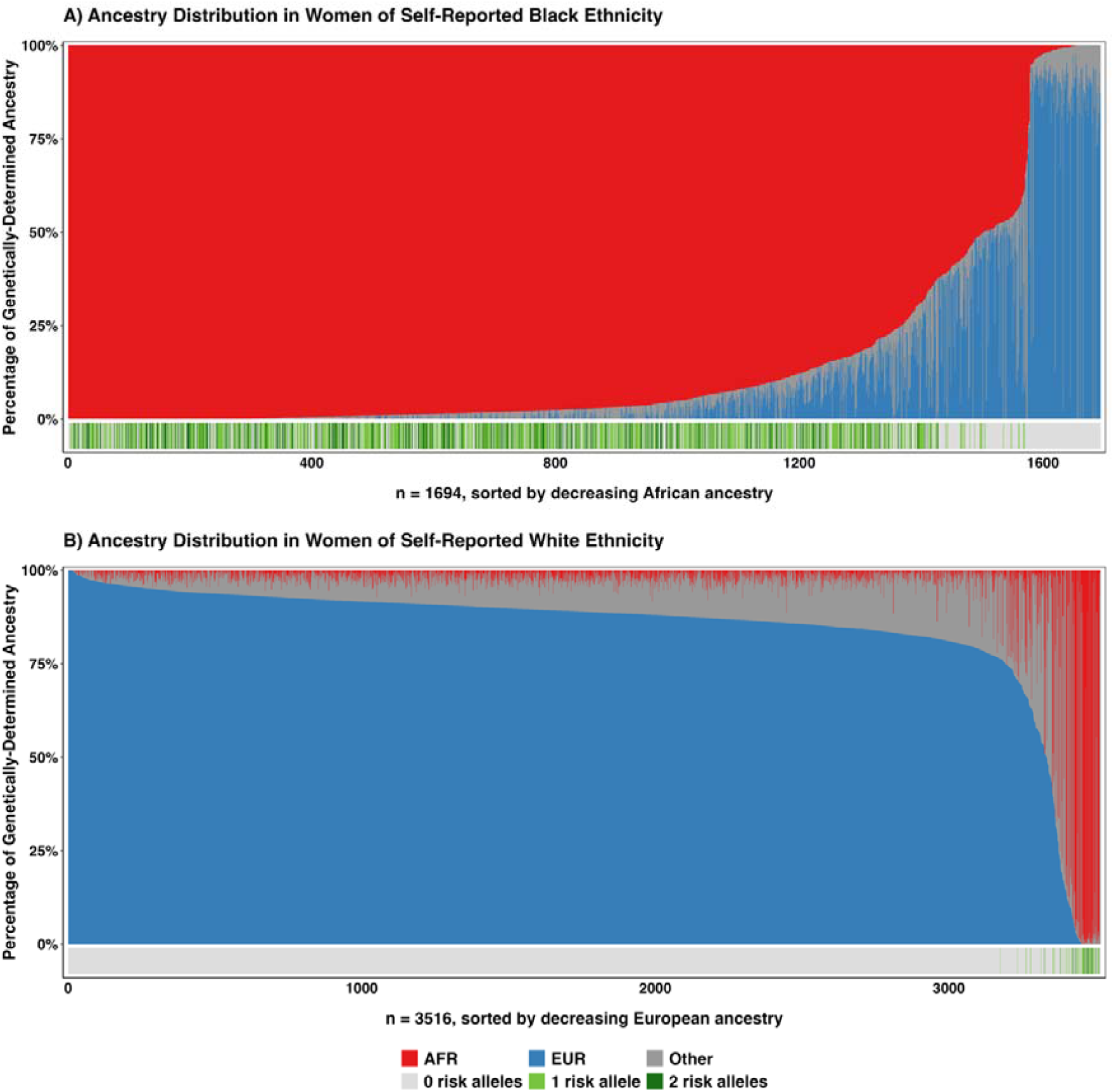
Distribution of *APOL1* alleles and Genetically-Determined Ancestry in Women of Self-Reported Black and White Ethnicity. Each woman is represented by a vertical bar divided into proportion of genetically-determined pan-African ancestry (AFR), European ancestry (EUR) and other ancestry (Other). Panel A shows women of self-reported Black ethnicity (n = 1694), sorted by decreasing African ancestry. Panel B shows women of self-reported White ethnicity (n = 3516), sorted by decreasing European ancestry. The horizontal strip below each panel indicates the number of *APOL1* risk alleles carried by each individual.

### Association of *APOL1* Risk Alleles with Preeclampsia Risk under a Recessive Model

The distribution *APOL1* risk alleles in women without preeclampsia and with preeclampsia stratified by SRE and GDA is shown in Figure S2.

After full clinical risk factor adjustment, the OR for preeclampsia risk in women with two *APOL1* risk alleles compared to one or zero risk alleles was 1.00 (95% CI 0.76 – 1.29) in the entire study population, 0.94 (95% CI 0.61 – 1.25) in women of Black SRE and 4.0 (95% CI 1.26 – 15.4) in women of White SRE (Figures 2A-C). Of the 16 women of White SRE with two *APOL1* risk alleles, 12 developed preeclampsia and four were controls (Figure S2A). Of these 16 women, only one woman had >50% EUR genetic ancestry, whilst 15 had >50% AFR genetic ancestry.

**Figure 2.**
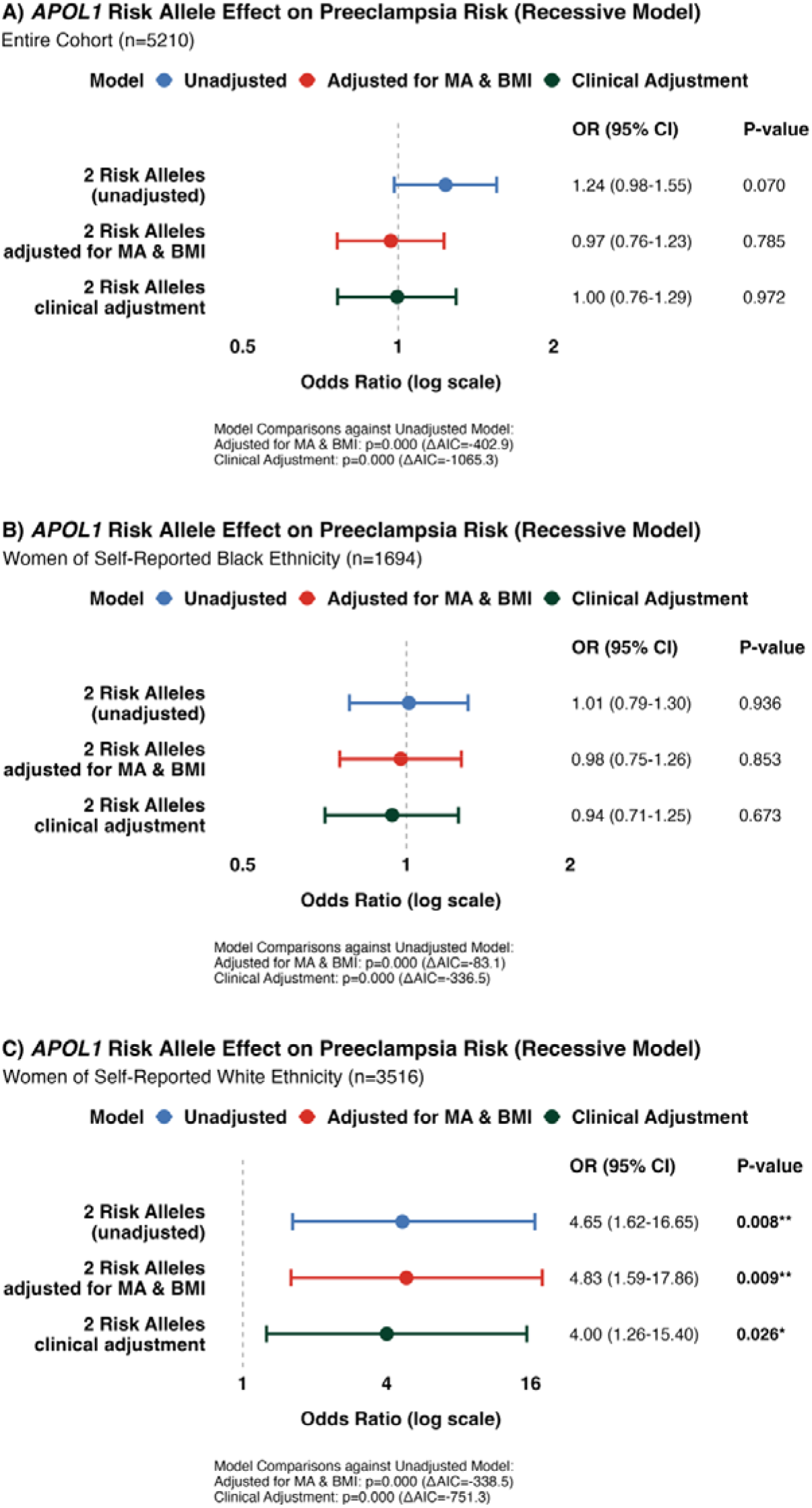
Association of *APOL1* Risk Alleles with Preeclampsia Risk under the Recessive Model. Forest plots demonstrating odds ratios for association between presence of two *APOL1* risk alleles (G1G1, G1G2 or G2G2) compared to zero or one risk alleles(G0G0, G1G0 or G2G0) and preeclampsia risk in the unadjusted model, model with adjustment for maternal age (MA) and body mass index (BMI) and model with clinical risk factor adjustment in the entire study population (A), in women of self-reported Black ethnic background (B) and self-reported White ethnic background (C). Model fit and comparison was assessed using likelihood ratio tests and the Akaike Information Criterion (AIC). OR = odds ratio, CI = confidence interval.

The OR for preeclampsia risk in women with >50% AFR genetic ancestry with two *APOL1* risk alleles compared to one or zero risk alleles was 1.00 (95% CI 0.76 – 1.32) after clinical risk factor adjustment (Figure S3A).

Regression analyses of preeclampsia risk in women with two *APOL1* risk alleles and one *APOL1* risk allele compared to zero *APOL1* risk alleles (additive inheritance model) and in women with one or two *APOL1* risk alleles compared to zero *APOL1* risk alleles (dominant inheritance model) have been included in Figures S4 and S5, respectively.

### Association of *APOL1* Risk Alleles with Early Preeclampsia Risk under the Recessive Model

Distribution of *APOL1* risk alleles in women without early preeclampsia and with early preeclampsia by SRE and GDA is shown in Figure S6.

The OR for early preeclampsia risk in women with two *APOL1* risk alleles compared to one or zero risk alleles in the entire study population was 1.34 (95% CI 0.83 – 2.06) after full clinical risk factor adjustment (Figure 3A). Similarly, no statistically significant associations were found between *APOL1* risk alleles in women of Black SRE (OR 1.03, 95% CI 0.63 – 1.63; Figure 3B) or women with >50% AFR genetic ancestry (OR 0.97, 95% CI 0.59 – 1.55; Figure S3B).

**Figure 3.**
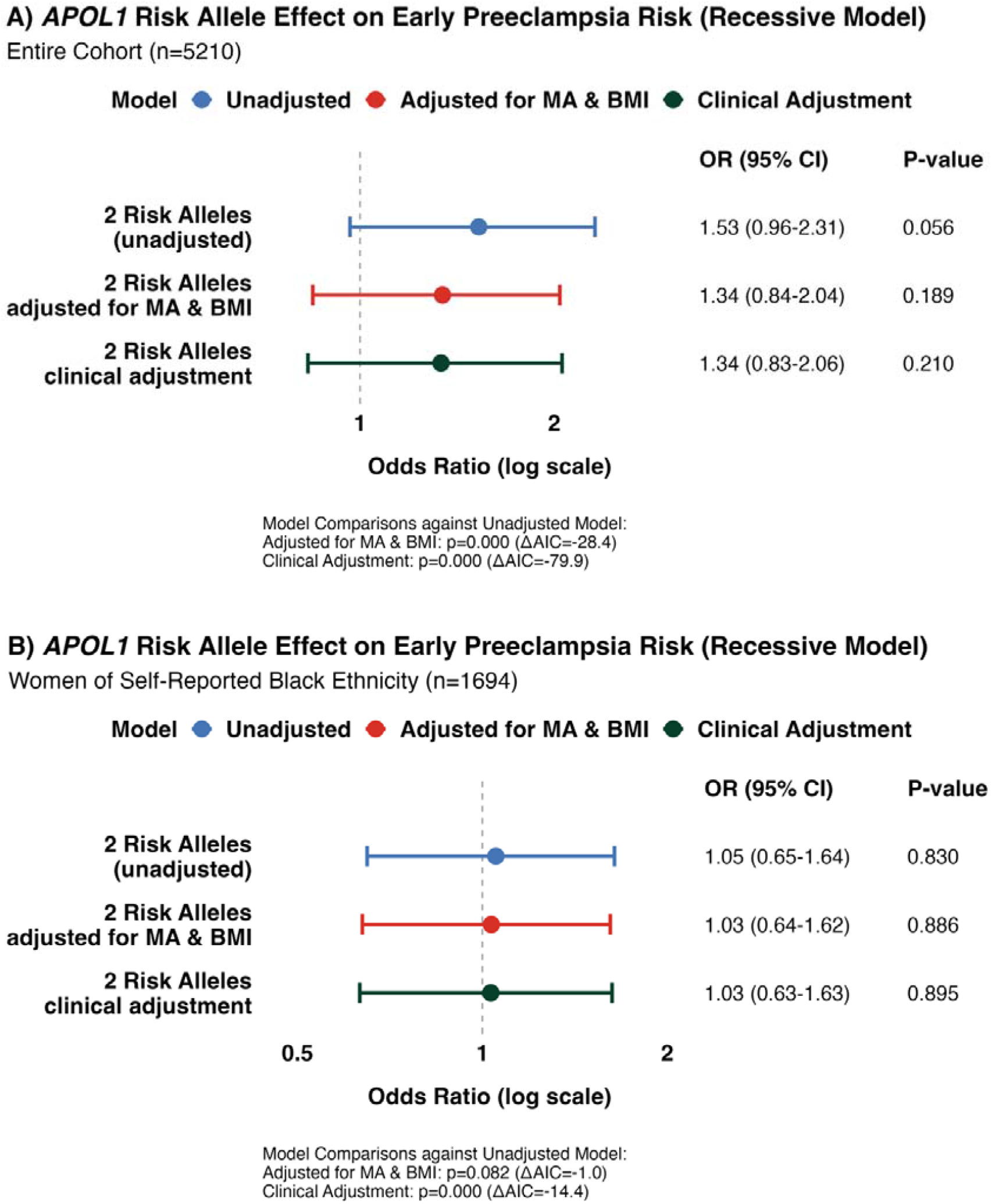
Association of *APOL1* Risk Alleles with Early Preeclampsia Risk under the Recessive Model. Forest plots demonstrating odds ratios for association between presence of two *APOL1* risk alleles (G1G1, G1G2 or G2G2) compared to zero or one risk alleles (G0G0, G1G0 or G2G0) and early preeclampsia risk in the unadjusted model, model with adjustment for maternal age (MA) and body mass index (BMI) and model with clinical risk factor adjustment in the entire study population (A) and in women of self-reported Black ethnic background (B). Model fit and comparison was assessed using likelihood ratio tests and the Akaike Information Criterion (AIC). OR = odds ratio, CI = confidence interval.

Additive and dominant inheritance model results are shown in Figures S7 and S8, respectively.

### Association of *APOL1* Risk Alleles with Recurrent Preeclampsia Risk under the Recessive Model

Distribution of *APOL1* risk alleles in multiparous women in the controls without previous preeclampsia and multiparous women in the preeclampsia cases with previous preeclampsia by SRE and GDA is shown in Figure S9.

In women with recurrent preeclampsia and multiparous controls, the OR for recurrent preeclampsia risk in women with two *APOL1* risk alleles was 0.57 (95% CI 0.27 – 1.10) after full adjustment (Figure 4A). Similar null associations were observed in women of Black SRE (OR 0.75, 95% CI 0.35 – 1.48; Figure 4B) and >50% AFR genetic ancestry subgroups (OR 0.74, 95% CI 0.34 – 1.45; Supplementary Figure S3C).

**Figure 4.**
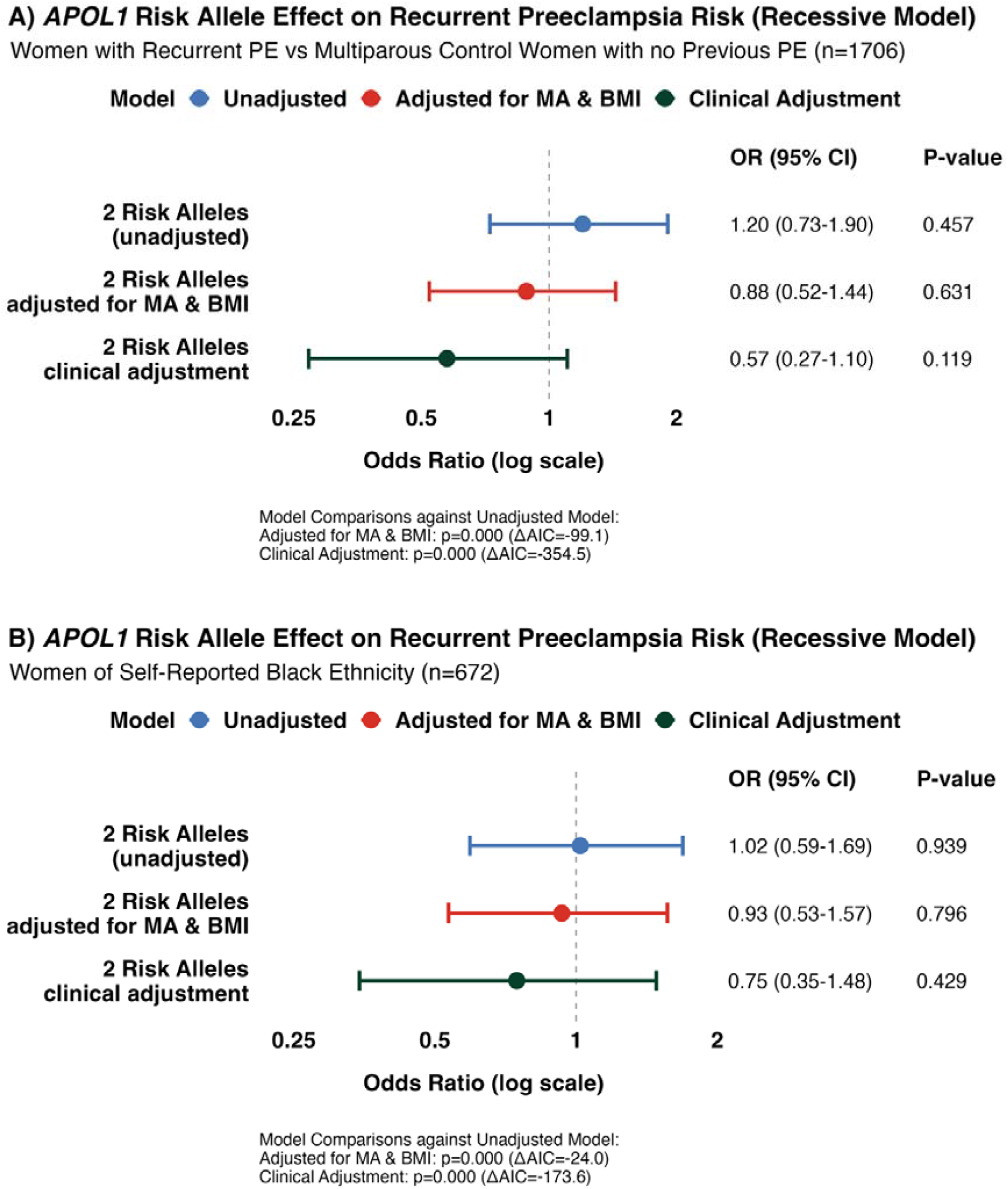
Association of *APOL1* Risk Alleles with Recurrent Preeclampsia Risk under the Recessive Model. Forest plots demonstrating odds ratios for association between presence of two *APOL1* risk alleles (G1G1, G1G2 or G2G2) compared to zero or one risk alleles (G0G0, G1G0 or G2G0) and recurrent preeclampsia (PE) risk in the unadjusted model, model with adjustment for maternal age (MA) and body mass index (BMI) and model with clinical risk factor adjustment in women with recurrent preeclampsia compared to multiparous women in the controls with no previous preeclampsia in the entire study population (A) and in women of self-reported Black ethnic background (B). Model fit and comparison was assessed using likelihood ratio tests and the Akaike Information Criterion (AIC). OR = odds ratio, CI = confidence interval.

Additive and dominant inheritance model results are shown in Figures S10 and S11, respectively.

### Association of *APOL1* Risk Alleles with Birthweight Centile and Gestational Age at Delivery

Distribution of birthweight centiles and gestational age at delivery in women with zero, one and two *APOL1* risk alleles in women without preeclampsia and with preeclampsia stratified by SRE and GDA is shown in Figures S12 and S13, respectively.

In controls, presence of two *APOL1* risk alleles was associated with lower birthweight centile (-6.9%, 95% CI -11.3% – -2.5%) after full adjustment (Figure S14A). This association varied by ethnicity, with no significant effect in women of Black SRE (1.0%, 95% CI -3.6% – 5.6%; Figure S14C) but a positive effect in women of White SRE (35.9%, 95% CI 8.8% – 63.0%; Figure S14E), though all four White women with two risk alleles had >50% AFR genetic ancestry. In control women with >50% AFR genetic ancestry, no significant association was observed (2.2%, 95% CI -2.5% – 6.8%; Figure S15A). In the preeclampsia cases, no significant associations were found between two *APOL1* risk alleles and birthweight centile across the entire study population (-3.9%, 95% CI -9.3% – 1.6%; Figure S14B) or ethnic subgroups (Figures S14D, S14F and S15B).

In the controls, no significant associations were observed between two *APOL1* risk alleles and gestational age at delivery across the entire study population (-0.06 weeks, 95% CI -0.34 – 0.21; Figure S16A) or ethnic subgroups (Figures S16C, S16E and S15C). In the preeclampsia cases, presence of two *APOL1* risk alleles was associated with earlier delivery (-0.56 weeks, 95% CI -1.08 – -0.04; Figure S16B). However, this association was not significant when stratified by ethnicity (Figures S16D, S16F and S15D).

## Comment

### Principal Findings

**In this large, nested case–control study of 5**,**210 multi-ethnic women, there was no evidence of association between maternal *APOL1* risk alleles and preeclampsia or related pregnancy outcomes**. Importantly, when restricting analysis to women of Black SRE or >50% AFR GDA only, the populations in whom maternal *APOL1* risk alleles are most relevant, no association was observed between maternal *APOL1* genotype and preeclampsia risk. An apparent association was observed in women of White SRE for preeclampsia risk in those with two *APOL1* risk alleles compared to one or zero risk alleles, which persisted after adjustment for clinical risk factors. However, genetic ancestry analysis revealed that 15 of the 16 women with White SRE who carried two *APOL1* risk alleles had >50% AFR GDA, and that genetically estimated ancestry was driving the association rather than the *APOL1* locus, as reported in previous studies[33].

Similarly, there was no evidence of independent associations between maternal *APOL1* risk alleles and birthweight centile or gestational age at delivery, with consistent findings in sensitivity analyses with stratification by SRE and GDA. These findings suggest that maternal *APOL1* risk alleles do not have an independent causal relationship with preeclampsia or associated outcomes like preterm birth or low birthweight. Associations observed in unadjusted analyses reflect the fact that *APOL1* risk alleles serve as a proxy for recent West African genetic ancestry.

### Strengths and Limitations

Our strengths include high-quality clinical phenotyping, evidenced by minimal missing data, comprehensive description of established preeclampsia risk factors, and preeclampsia diagnoses made by trained clinicians based on international guidelines[37].

Furthermore, our current understanding of the genetic architecture of preeclampsia across diverse populations is Eurocentric and limited by modest sample sizes[26,38-42], and this is the largest genetic study of maternal *APOL1* renal risk alleles and preeclampsia in women of Black ethnic backgrounds[24,26]. Thus, our study provides diversification of genetic datasets necessary to enable genomic and molecular insights to benefit women with the highest burden of preeclampsia[43,44]. However, we may not have had enough power to detect weaker associations, particularly when examining extreme preeclampsia phenotypes.

A further strength of our study was the ability to enhance maternal SRE data with individual GDA, which enabled deeper understanding of the observed association between *APOL1* alleles and disease risk in White women. It has been previously shown that self-reported ethnicity is an imperfect proxy for genetic ancestry, particularly in individuals of mixed ancestry[33,45].

### Results in the Context of What is Known

#### 1. Fetal *APOL1* Genotype

Several small case and case-control studies have reported associations between fetal *APOL1* risk allele status and preeclampsia risk, rather than maternal *APOL1* risk allele status alone[24,46]. Two studies demonstrated that fetal high-risk genotypes (two *APOL1* risk alleles) were associated with increased preeclampsia risk (OR 1.84, 95% CI 1.11 – 2.93 and OR 1.92, 95% CI 1.05 – 3.49 in cohorts of n=121 and n=93 cases, respectively), whilst maternal *APOL1* genotypes showed no association[24]. Further investigations have suggested that maternal-fetal *APOL1* genotype discordance may confer additional risk, with one study reporting a 2.6-fold increased preeclampsia risk when maternal and fetal genotypes differed[26]. It is possible that smaller studies reporting associations between maternal *APOL1* status and preeclampsia risk may have inadvertently captured fetal genetic effects due to parent-offspring genetic correlation, leading to spurious associations.

#### 2. Mode of inheritance

Whilst most studies to date suggest a recessive model for *APOL1* risk alleles, some studies have suggested dominant or additive inheritance patterns[25,26,46-48]. However, in our study, we found no evidence for association between *APOL1* variants and preeclampsia under recessive, dominant, or additive inheritance models. This strengthens our conclusion that maternal *APOL1* variants do not meaningfully contribute to preeclampsia risk in our population, regardless of assumed mode of inheritance.

#### 3. APOL1-Environment Interaction

The lack of association between maternal *APOL1* genotype with preeclampsia risk and preeclampsia-associated outcomes in our study could be consistent with emerging evidence surrounding gene-environment interactions, with a ‘second hit’ being required for clinical phenotype manifestation[49]. Polygenic variants may also modify the impact of rare variants[50,51]. *APOL1* risk allele expression is interferon-inducible, with podocyte death and glomerular scarring potentiated by a hyperinflammatory state, such as viral infections or autoimmune conditions[49,52-55]. It is plausible that the inflammatory milieu characteristic of preeclampsia could serve as the trigger to initiating pathways that contribute to long-term maternal cardiovascular and renal complications associated with *APOL1* risk alleles without immediate adverse outcomes[56].

It has also been suggested that prolonged exposure to environmental conditions, such as chronic stress or social adversity, may influence whether *APOL1* genotype confers preeclampsia risk[23]. In a study of 426 women, Hong et al. showed that, whilst *APOL1* risk genotypes were associated with preeclampsia in African American women, this was not observed in Haitian women living in the US, even though both groups shared similar West African ancestry[26]. Factors such as universal healthcare access in the UK, inclusion in a research study with personalized risk assessment and recommendation for aspirin prophylaxis, may have affected whether maternal *APOL1* risk alleles conferred preeclampsia risk in the context of this study, consistent with our emerging understanding of gene-environment interactions in multifactorial diseases including preeclampsia[57].

### Clinical Implications

Our study found no evidence of association between maternal *APOL1* genotype and pre-eclampsia risk. Our data suggests the apparent association between *APOL1* risk alleles and preeclampsia reflects West African genetic ancestry as a proxy rather than a direct causal effect of the *APOL1* locus. This suggests *APOL1* genotype should not currently be incorporated into preeclampsia screening algorithms.

### Research Implications

First, larger studies examining both maternal and fetal *APOL1* genotypes simultaneously are needed to disentangle their respective contributions to preeclampsia risk, given evidence that fetal high-risk genotypes and maternal-fetal genotype discordance may be more important than maternal genotype alone. Second, longitudinal studies examining maternal outcomes in women with *APOL1* risk alleles and preeclampsia are needed to establish whether preeclampsia-associated inflammation acts as a second hit to activate *APOL1*-mediated organ damage. Finally, expanding genetic studies of preeclampsia to include larger, more diverse populations with comprehensive ancestry data will be essential to overcome the Eurocentric bias that currently limits our understanding of the genetic architecture of preeclampsia, with exploration of polygenic contribution to disease as well as rare disease loci such as *APOL1*.

## Conclusions

We found no evidence of an independent association between maternal *APOL1* risk alleles and preeclampsia or related adverse outcomes (preterm birth, low birthweight) in a multi-ethnic UK population. Our findings suggest that *APOL1* alleles serve as a proxy for West African genetic ancestry rather than having a direct causal relationship with preeclampsia.

## Data Availability

All data produced in the present study are available upon reasonable request to the authors

## Declaration of Interests

FCR receives part-time salary contribution as Chief Medical Officer at MEGI Health UK Ltd and receives consulting fees for advisory services provided through Option 5 Health Limited, Revena Limited and Gerson Lehrman Group Limited (GLG). The remaining authors have nothing to disclose.

An abstract derived from this paper has been accepted for oral presentation at the 10^th^ International Congress on Cardiac Problems in Pregnancy 16-19^th^ April in Barcelona, Spain

## Glossary

AIC: Akaike information criterion
CI: Confidence interval
IVF: In vitro fertilization
SRE: Self-reported ethnicity
BMI: Body mass index
GDA: Genetically-determined ancestry
OR: Odds ratio

## Author Contributions

*Wen Tong:* Methodology, Software, Validation, Formal analysis, Data curation, Writing – Original Draft, Writing – Review & Editing, Visualization

*Frances Conti-Ramsden:* Conceptualization, Methodology, Software, Validation, Investigation, Resources, Data curation, Writing – Original Draft, Writing – Review & Editing, Supervision, Project administration

*Hannah Beckwith:* Conceptualization, Methodology, Software, Validation, Investigation, Resources, Data curation, Writing – Original Draft, Writing – Review & Editing, Supervision, Project administration

*Argyro Syngelaki:* Resources, Data curation, Writing – Review & Editing

Ioannis Mitrogiannis: Resources, Data curation

*Lucy Chappell:* Conceptualization, Methodology, Writing – Review and Editing, Supervision Pirro

*Hysi:* Methodology, Software, Data cleaning, Data analysis

*Catherine Williamson:* Conceptualization, Methodology, Investigation, Resources, Writing – Review & Editing, Supervision, Funding acquisition

*Sophie Limou:* Methodology, Validation, Review & Editing, Supervision

*Kypros Nicolaides:* Conceptualization, Methodology, Resources, Writing – Review & Editing, Supervision, Project administration, Funding acquisition

*Kate Bramham:* Conceptualization, Methodology, Resources, Writing – Review & Editing, Supervision, Project administration, Funding acquisition

*Antonio de Marvao:* Conceptualization, Methodology, Resources, Writing – Review & Editing, Supervision, Project administration, Funding acquisition

## Supplementary Material

### Supplementary Tables

**Supplementary Table S1.** Baseline Characteristics, Clinical Risk Factors for Preeclampsia, *APOL1* Risk Allele Distribution in Women with Preeclampsia and without Preeclampsia Stratified by Genetically-Determined Ancestry. Results are presented as mean (standard deviation) or median [interquartile range] for continuous variables and count (percentage) for categorical variables. Abbreviations: AFR = genetically-determined pan-African ancestry, EUR = genetically-determined European ancestry, PE = preeclampsia; Definitions: Early preeclampsia = delivery with preeclampsia before 34 weeks gestational age, preterm preeclampsia = delivery with preeclampsia between 34 and 37 weeks gestational age, term preeclampsia = delivery with preeclampsia after 37 weeks gestational age.

| *Genetically-Determined Ancestry* | **>50% AFR**<br>1599 (30.7%) |  | **>50% EUR**<br>3464 (66.5%) |  | **Admixed**<br>147 (2.8%) |  |
| --- | --- | --- | --- | --- | --- | --- |
| *Study Group* | **Preeclampsia** | **Control** | **Preeclampsia** | **Control** | **Preeclampsia** | **Control** |
| **Number Of Women** | 708 (44.3%) | 891 (55.7%) | 1377 (39.8%) | 2087 (60.2%) | 45 (30.6%) | 102 (69.4%) |
| - *Early Preeclampsia* | 106 (15.0%) |  | 130 (9.4%) |  | 3 (6.7%) |  |
| - *Preterm Preeclampsia* | 111 (15.7%) |  | 161 (11.7%) |  | 6 (13.3%) |  |
| - *Term Preeclampsia* | 491 (69.4%) |  | 1086 (78.9%) |  | 36 (80.0%) |  |
| **Baseline Maternal Characteristics** |  |  |  |  |  |  |
| **Age (years)** | 31.5 ± 6.2 | 31.1 ± 5.5 | 31.8 ± 5.8 | 33.2 ± 4.6 | 31.7 ± 6.3 | 31.9 ± 5.5 |
| **Body Mass Index (kg/m<sup>2</sup>)** | 30.2 ± 6.7 | 27.7 ± 5.6 | 28.1 ± 6.5 | 24.6 ± 4.4 | 27.0 ± 5.8 | 25.6 ± 4.6 |
| **Chronic Hypertension** | 128 (18.1%) | 6 (0.7%) | 100 (7.3%) | 1 (0.0%) | 4 (8.0%) | 0 (0.0%) |
| **Diabetes Mellitus** |  |  |  |  |  |  |
| - *Type 1 Diabetes* | 5 (0.7%) | 2 (0.2%) | 20 (1.5%) | 4 (0.2%) | 0 (0.0%) | 0 (0.0%) |
| - *Type 2 Diabetes* | 21 (3.0%) | 22 (2.5%) | 11 (0.8%) | 20 (1.0%) | 3 (6.0%) | 1 (1.0%) |
| **Assisted Pregnancy** |  |  |  |  |  |  |
| - *Ovulation Induction* | 12 (1.7%) | 1 (0.1%) | 17 (1.2%) | 19 (0.9%) | 0 (0.0%) | 0 (0.0%) |
| - *In Vitro Fertilization* | 16 (2.3%) | 11 (1.2%) | 99 (7.2%) | 145 (6.9%) | 1 (2.0%) | 6 (5.9%) |
| **Smoking** | 12 (1.7%) | 21 (2.4%) | 80 (5.8%) | 48 (2.3%) | 0 (0.0%) | 1 (1.0%) |
| **Parity** |  |  |  |  |  |  |
| - *Nulliparous* | 336 (47.5%) | 352 (39.5%) | 950 (69.0%) | 1183 (56.7%) | 27 (60.0%) | 47 (46.1%) |
| - *Multiparous – No PE* | 264 (37.3%) | 516 (57.9%) | 283 (20.6%) | 873 (41.8%) | 7 (15.0%) | 54 (52.9%) |
| - *Recurrent PE* | 108 (15.3%) | 23 (2.6%) | 144 (10.5%) | 31 (1.5%) | 11 (24.0%) | 1 (1.0%) |
| **Family History of PE** | 68 (9.6%) | 53 (5.9%) | 150 (10.9%) | 80 (3.8%) | 2 (4.0%) | 1 (1.0%) |
| **Pregnancy Outcomes** |  |  |  |  |  |  |
| **Birthweight (g)** | 2715 ± 888 | 3261 ± 534 | 2976 ± 799 | 3436 ± 513 | 2911 ± 672 | 3360 ± 534 |
| **Birthweight Centile (%)** | 30.0 ± 31.4 | 41.5 ± 28.3 | 37.9 ± 32.3 | 51.2 ± 28.4 | 33.5 ± 28.3 | 48.6 ± 29.2 |
| **Gestational Age at Delivery (weeks)** | 38.1 [36.3-39.6] | 39.9 [39.0-40.6] | 38.7 [37.3-40.0] | 40.0 [39.1-40.9] | 38.6 [37.3-40.1] | 40.0 [38.8-40.6] |
| **Stillbirth** | 18 (2.5%) | 2 (2.2%) | 15 (1.1%) | 2 (0.1%) | 0 (0.0%) | 0 (0.0%) |
| **Neonatal Death** | 0 (0.0%) | 0 (0.0%) | 2 (0.1%) | 2 (0.1%) | 0 (0.0%) | 0 (0.0%) |
| **APOL1 Risk Allele Frequency (%)** |  |  |  |  |  |  |
| *rs73885319* (G1<sup>G</sup>) | 339 (47.9%) | 414 (46.0%) | 3 (0.2%) | 2 (0.1%) | 1 (2.2%) | 6 (5.9%) |
| *rs60910145* (G1<sup>M</sup>) | 339 (47.9%) | 414 (46.0%) | 3 (0.2%) | 2 (0.1%) | 1 (2.2%) | 6 (5.9%) |
| *rs73885319* or *rs60910145* (G1) | 339 (47.9%) | 414 (46.0%) | 3 (0.2%) | 2 (0.1%) | 1 (2.2%) | 6 (5.9%) |
| *rs71785313* (G2) | 161 (22.7%) | 237 (26.6%) | 2 (0.1%) | 2 (0.1%) | 1 (2.2%) | 2 (2.0%) |
| **1 APOL1 Risk Allele** | 303 (42.8%) | 410 (46.0%) | 3 (0.2%) | 4 (0.2%) | 2 (4.4%) | 8 (7.8%) |
| **2 APOL1 Risk Alleles** | 144 (20.3%) | 172 (19.3%) | 1 (0.1%) | 0 (0.0%) | 0 (0.0%) | 0 (0.0%) |
| **APOL1 Genotype Frequency (%)** |  |  |  |  |  |  |
| **G1G1** | 59 (8.3%) | 84 (8.9%) | 0 (0.0%) | 0 (0.0%) | 0 (0.0%) | 0 (0.0%) |
| **G2G2** | 12 (1.7%) | 19 (2.1%) | 0 (0.0%) | 0 (0.0%) | 0 (0.0%) | 0 (0.0%) |
| **G1G2** | 73 (10.3%) | 69 (7.7%) | 1 (0.1%) | 0 (0.0%) | 0 (0.0%) | 0 (0.0%) |
| **G1G0** | 207 (29.2%) | 261 (29.0%) | 2 (0.1%) | 2 (0.1%) | 1 (2.2%) | 6 (5.9%) |
| **G2G0** | 96 (13.6%) | 149 (16.7%) | 1 (0.1%) | 2 (0.1%) | 1 (2.2%) | 2 (2.0%) |
| **G0G0** | 260 (36.7%) | 309 (34.7%) | 1373 (99.7%) | 2083 (99.8%) | 43 (95.6%) | 94 (92.2%) |

### Supplementary Figures

**Supplementary Figure S1.**
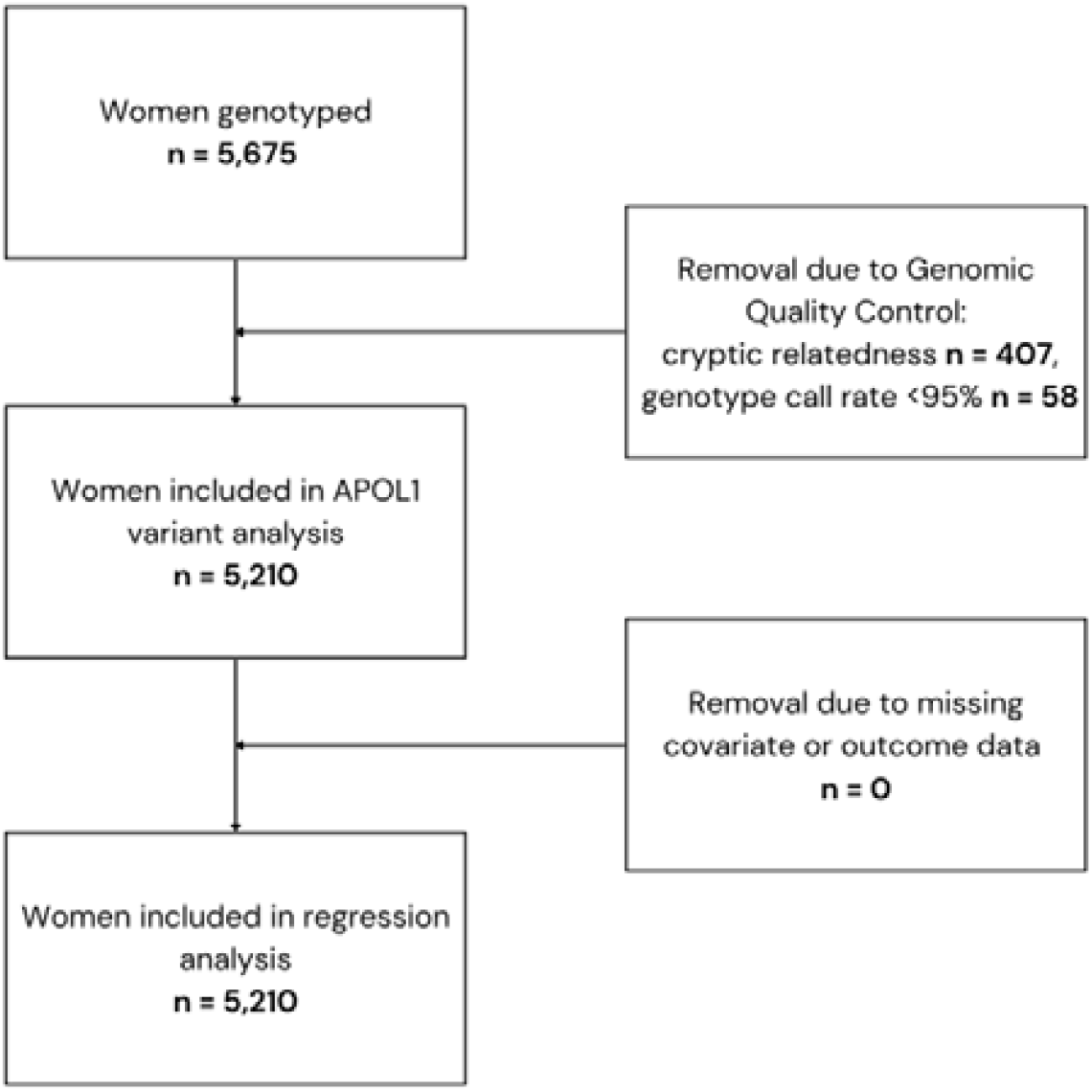
Study Flow Diagram

**Supplementary Figure S2.**
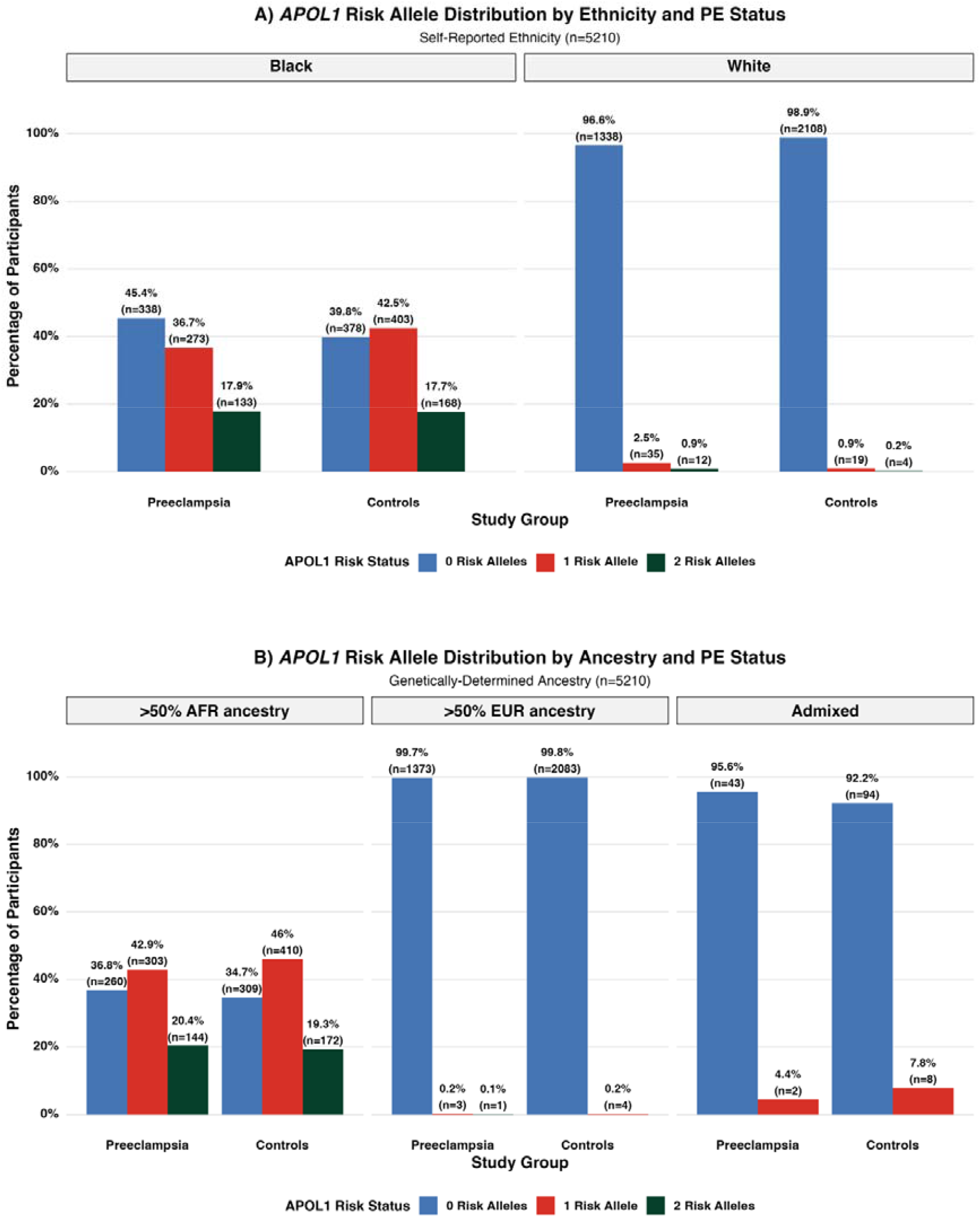
Distribution of *APOL1* Risk Alleles in Women with Preeclampsia and without Preeclampsia by Self-Reported Ethnicity (A) and by Genetically-Determined Ancestry (B) Abbreviations: PE = preeclampsia, AFR = genetically-determined pan-African ancestry, EUR = genetically-determined European ancestry.

**Supplementary Figure S3.**
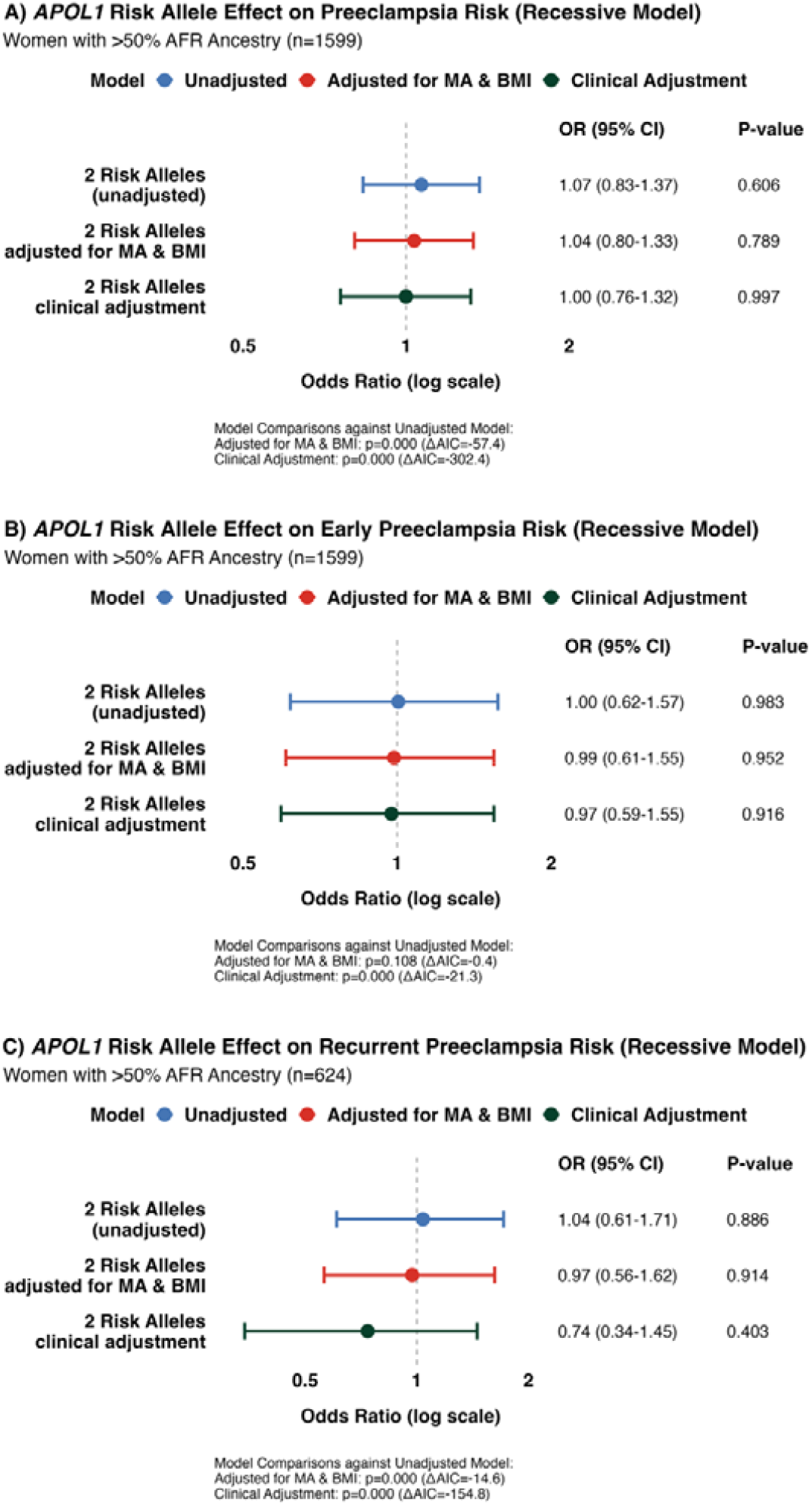
Association of *APOL1* Risk Alleles with Preeclampsia Risk, Early Preeclampsia Risk and Recurrent Preeclampsia Risk by Genetically-Determined Ancestry under the Recessive Model. Forest plots demonstrating odds ratios for association between presence of two *APOL1* risk alleles (G1G1, G1G2 or G2G2) compared to zero or one risk alleles (G0G0, G1G0 or G2G0) and preeclampsia risk (A), early preeclampsia risk (B) and recurrent preeclampsia risk (C) in the unadjusted model, model with adjustment for maternal age (MA) and body mass index (BMI) and model with clinical risk factor adjustment in women with >50% pan-African (AFR) genetic ancestry. Model fit and comparison was assessed using likelihood ratio tests and the Akaike Information Criterion (AIC). OR = odds ratio, CI = confidence interval; Definitions: Early preeclampsia = delivery with preeclampsia before 34 weeks gestational age, recurrent preeclampsia = women with preeclampsia with previous history of preeclampsia.

**Supplementary Figure S4.**
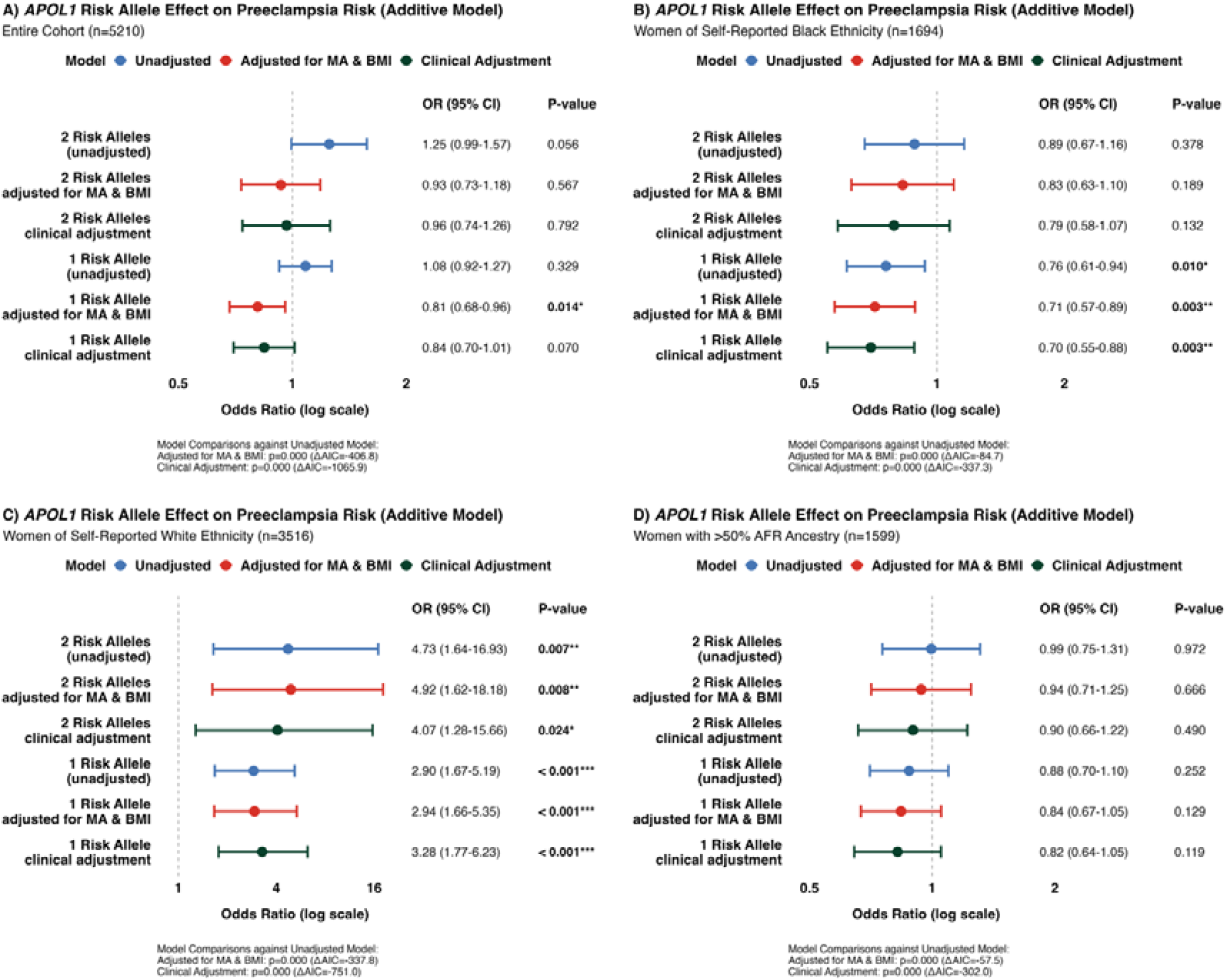
Association of *APOL1* Risk Alleles with Preeclampsia Risk under the Additive Model. Forest plots demonstrating odds ratios for association between presence of two *APOL1* risk alleles (G1G1, G1G2 or G2G2) and one *APOL1* risk allele (G1G0 or G2G0) compared to zero risk alleles (G0G0) and preeclampsia risk in the unadjusted model, model with adjustment for maternal age (MA) and body mass index (BMI) and model with clinical risk factor adjustment in the entire study population (A), in women of self-reported Black ethnic background (B), in women of self-reported White ethnic background (C) and in women with >50% pan-African (AFR) genetic ancestry (D). Model fit and comparison was assessed using likelihood ratio tests and the Akaike Information Criterion (AIC). OR = odds ratio, CI = confidence interval.

**Supplementary Figure S5.**
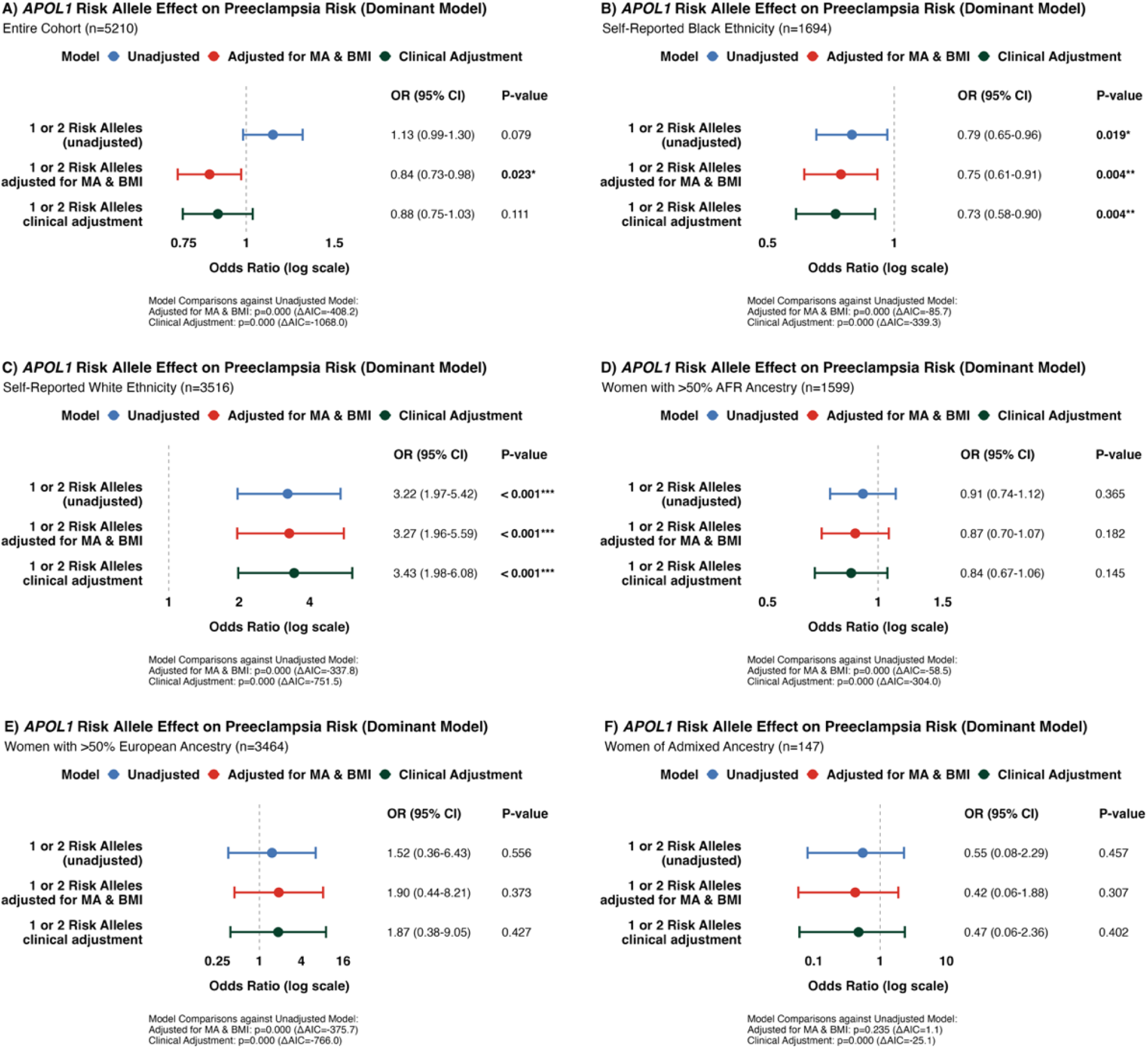
Association of *APOL1* Risk Alleles with Preeclampsia Risk under the Dominant Model. Forest plots demonstrating odds ratios for association between presence of two or one *APOL1* risk alleles (G1G1, G1G2, G2G2, G1G0 or G2G0) compared to zero risk alleles (G0G0) and preeclampsia risk in the unadjusted model, model with adjustment for maternal age (MA) and body mass index (BMI) and model with clinical risk factor adjustment in the entire study population (A), in women of self-reported Black ethnic background (B), in women of self-reported White ethnic background (C), in women with >50% pan-African (AFR) genetic ancestry (D), in women with >50% European (EUR) genetic ancestry (E) and in women with Admixed genetic ancestry (F). Model fit and comparison was assessed using likelihood ratio tests and the Akaike Information Criterion (AIC). OR = odds ratio, CI = confidence interval.

**Supplementary Figure S6.**
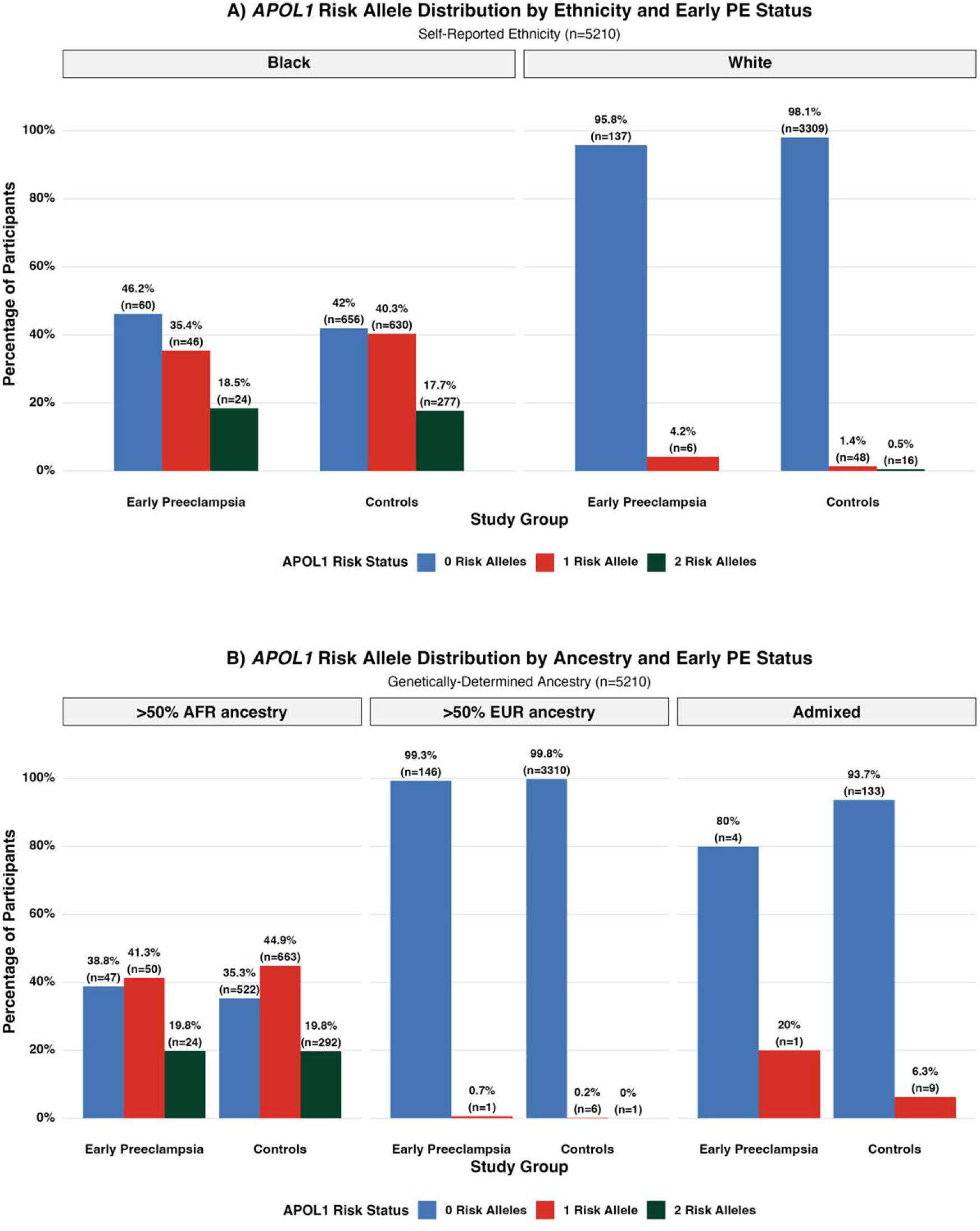
Distribution of *APOL1* Risk Alleles in Women with Early Preeclampsia and without Early Preeclampsia by Self-Reported Ethnicity (A) and by Genetically-Determined Ancestry (B) Abbreviations: PE = preeclampsia, AFR = genetically-determined pan-African ancestry, EUR = genetically-determined European ancestry; Definitions: Early preeclampsia = delivery with preeclampsia before 34 weeks gestational age

**Supplementary Figure S7.**
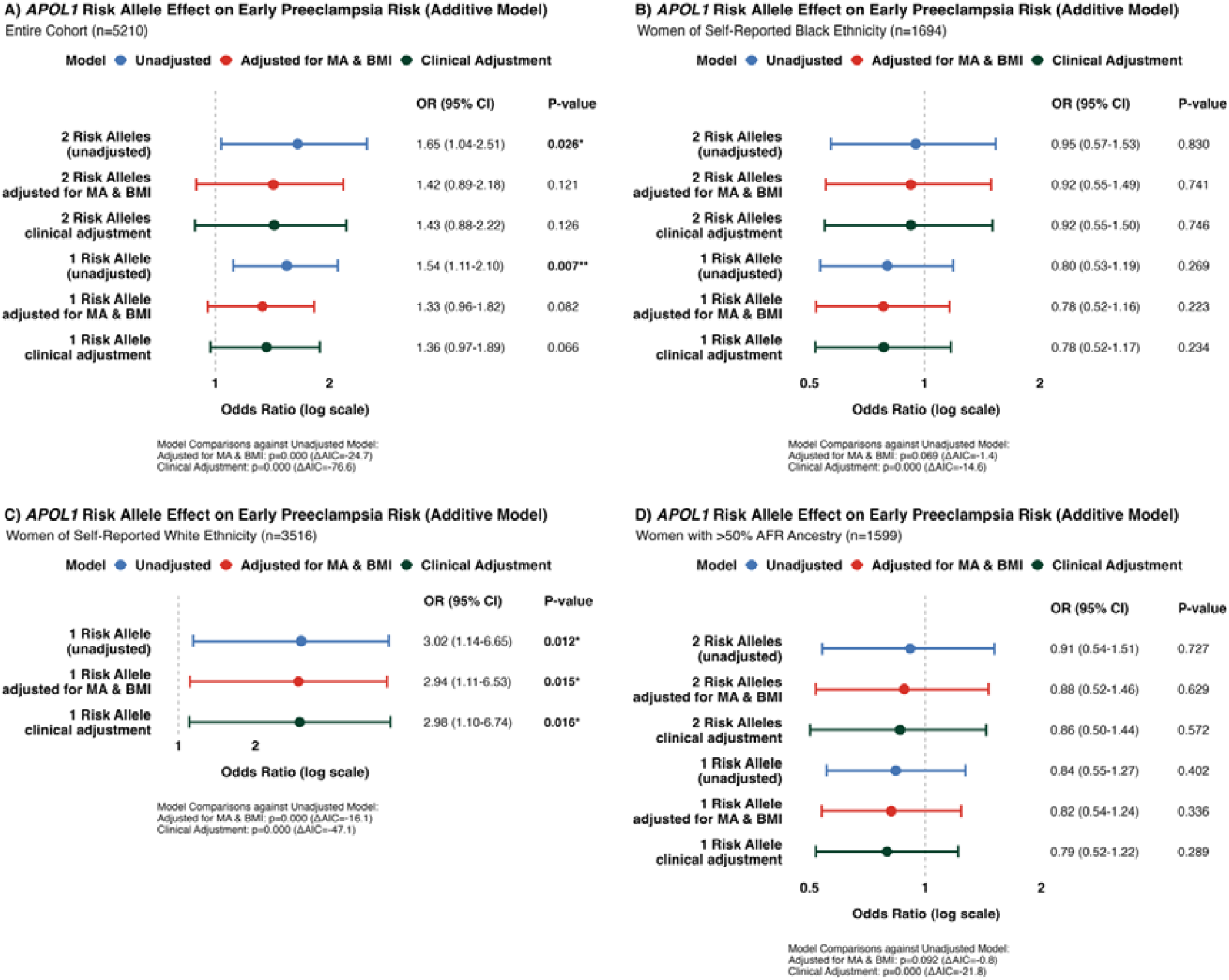
Association of *APOL1* Risk Alleles with Early Preeclampsia Risk under the Additive Model. Forest plots demonstrating odds ratios for association between presence of two *APOL1* risk alleles (G1G1, G1G2 or G2G2) and one *APOL1* risk allele (G1G0 or G2G0) compared to zero risk alleles (G0G0) and early preeclampsia risk in the unadjusted model, model with adjustment for maternal age (MA) and body mass index (BMI) and model with clinical risk factor adjustment in the entire study population (A), in women of self-reported Black ethnic background (B) in women of self-reported White ethnic background (C) and in women with >50% pan-African (AFR) genetic ancestry (D). Model fit and comparison was assessed using likelihood ratio tests and the Akaike Information Criterion (AIC). OR = odds ratio, CI = confidence interval.

**Supplementary Figure S8.**
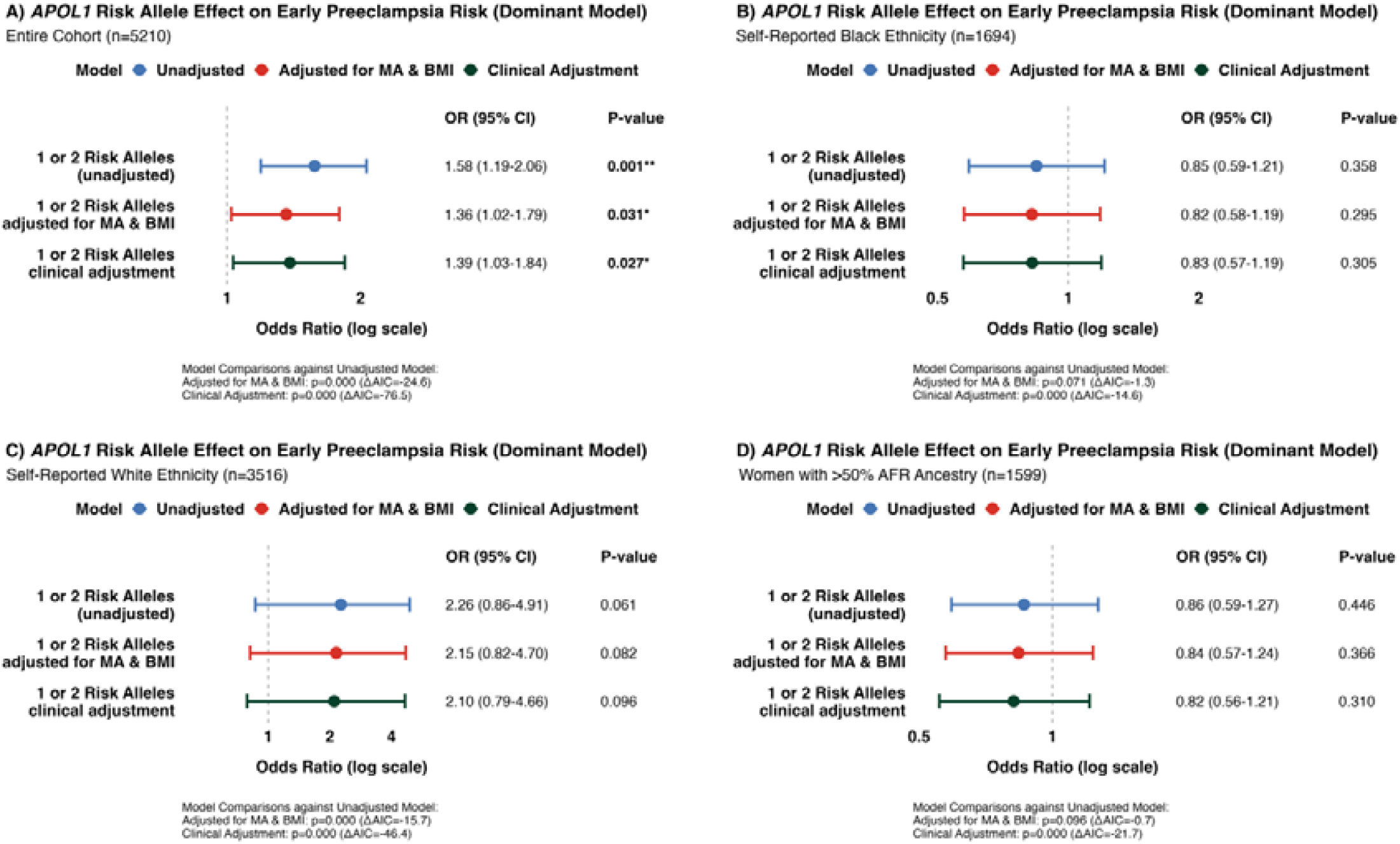
Association of *APOL1* Risk Alleles with Early Preeclampsia Risk under the Dominant Model. Forest plots demonstrating odds ratios for association between presence of two or one *APOL1* risk alleles (G1G1, G1G2, G2G2, G1G0 or G2G0) compared to zero risk alleles (G0G0) and early preeclampsia risk in the unadjusted model, model with adjustment for maternal age (MA) and body mass index (BMI) and model with clinical risk factor adjustment in the entire study population (A), in women of self-reported Black ethnic background (B) in women of self-reported White ethnic background (C) and in women with >50% pan-African (AFR) genetic ancestry (D). Model fit and comparison was assessed using likelihood ratio tests and the Akaike Information Criterion (AIC). OR = odds ratio, CI = confidence interval.

**Supplementary Figure S9.**
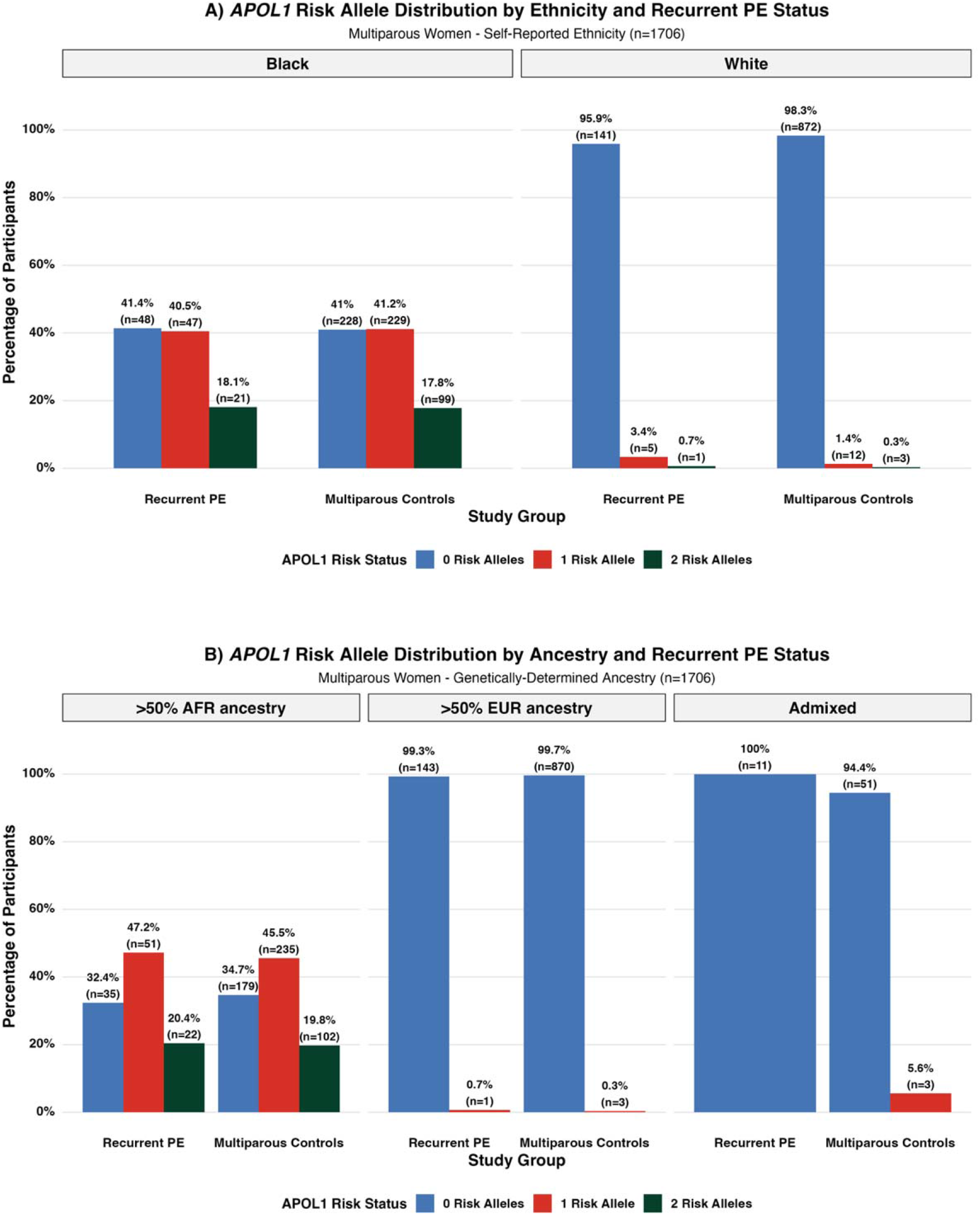
Distribution of *APOL1* Risk Alleles in Women with Recurrent Preeclampsia and in Multiparous Controls by Self-Reported Ethnicity (A) and by Genetically-Determined Ancestry (B) Abbreviations: PE = preeclampsia, AFR = genetically-determined pan-African ancestry, EUR = genetically-determined European ancestry; Definitions: recurrent preeclampsia = women with preeclampsia with previous history of preeclampsia, multiparous controls = multiparous women without preeclampsia with no previous history of preeclampsia

**Supplementary Figure S10.**
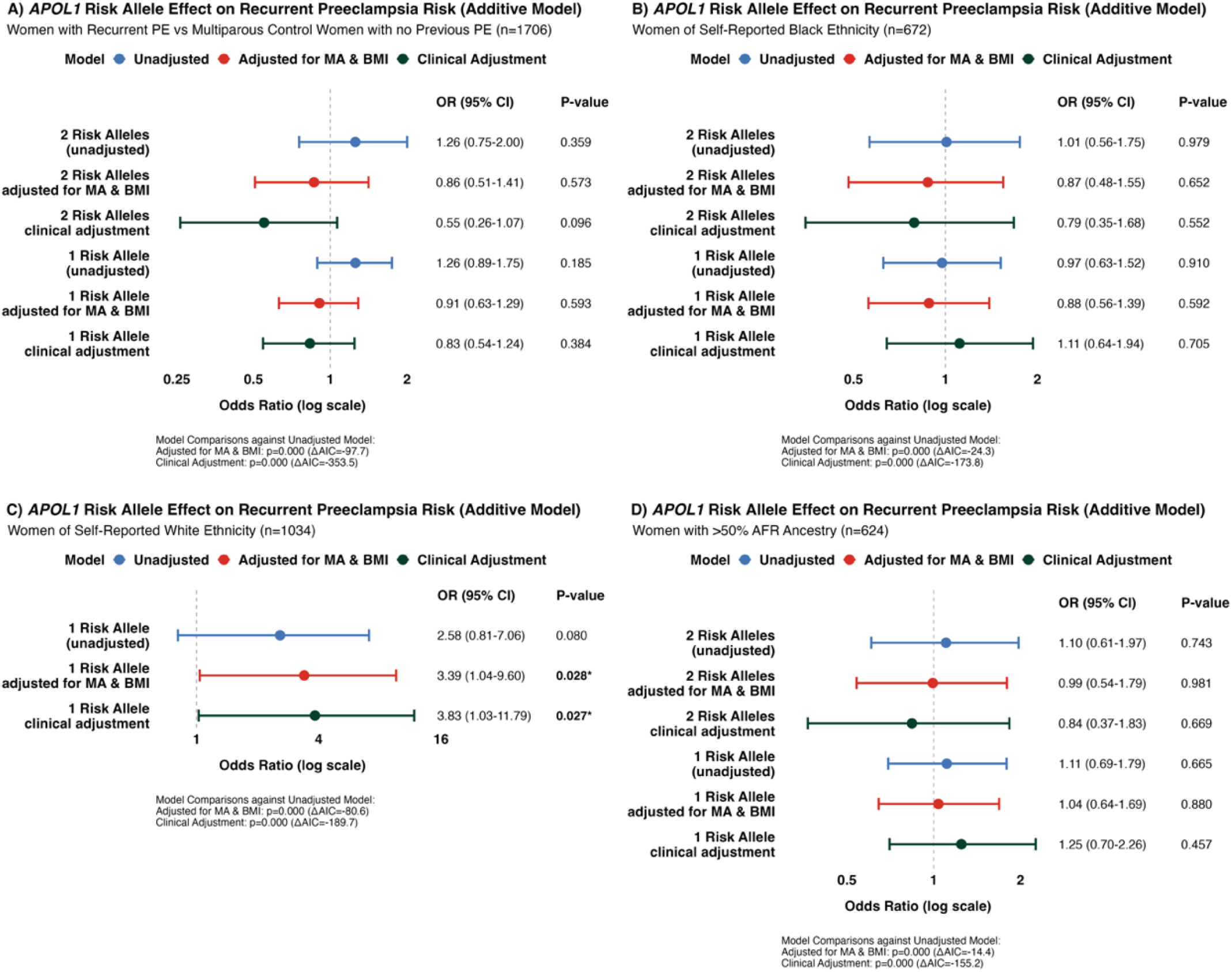
Association of *APOL1* Risk Alleles with Recurrent Preeclampsia Risk under the Additive Model. Forest plots demonstrating odds ratios for association between presence of two *APOL1* risk alleles (G1G1, G1G2 or G2G2) and one *APOL1* risk allele (G1G0 or G2G0) compared to zero risk alleles (G0G0) and recurrent preeclampsia (PE) risk in the unadjusted model, model with adjustment for maternal age (MA) and body mass index (BMI) and model with clinical risk factor adjustment in women with recurrent preeclampsia compared to multiparous women in the controls with no previous preeclampsia in the entire study population (A), in women of self-reported Black ethnic background (B), in women of self-reported White ethnic background (C) and in women with >50% pan-African (AFR) genetic ancestry (D). Model fit and comparison was assessed using likelihood ratio tests and the Akaike Information Criterion (AIC). OR = odds ratio, CI = confidence interval.

**Supplementary Figure S11.**
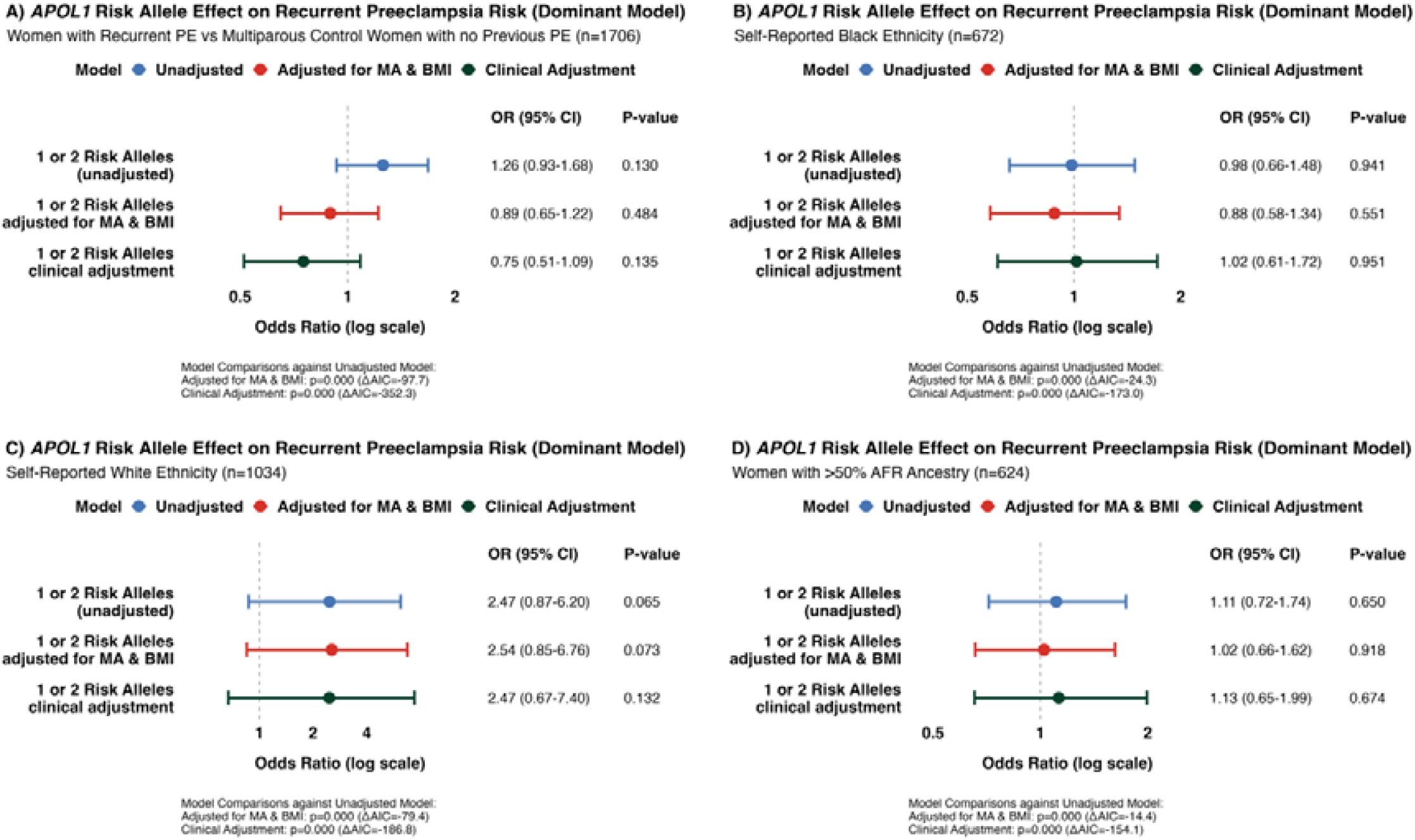
Association of *APOL1* Risk Alleles with Recurrent Preeclampsia Risk under the Dominant Model. Forest plots demonstrating odds ratios for association between presence of two or one *APOL1* risk alleles (G1G1, G1G2, G2G2, G1G0 or G2G0) compared to zero risk alleles (G0G0) and recurrent preeclampsia (PE) risk in the unadjusted model, model with adjustment for maternal age (MA) and body mass index (BMI) and model with clinical risk factor adjustment in women with recurrent preeclampsia compared to multiparous women in the controls with no previous preeclampsia in the entire study population (A), in women of self-reported Black ethnic background (B), in women of self-reported White ethnic background (C) and in women with >50% pan-African (AFR) genetic ancestry (D). Model fit and comparison was assessed using likelihood ratio tests and the Akaike Information Criterion (AIC). OR = odds ratio, CI = confidence interval.

**Supplementary Figure S12.**
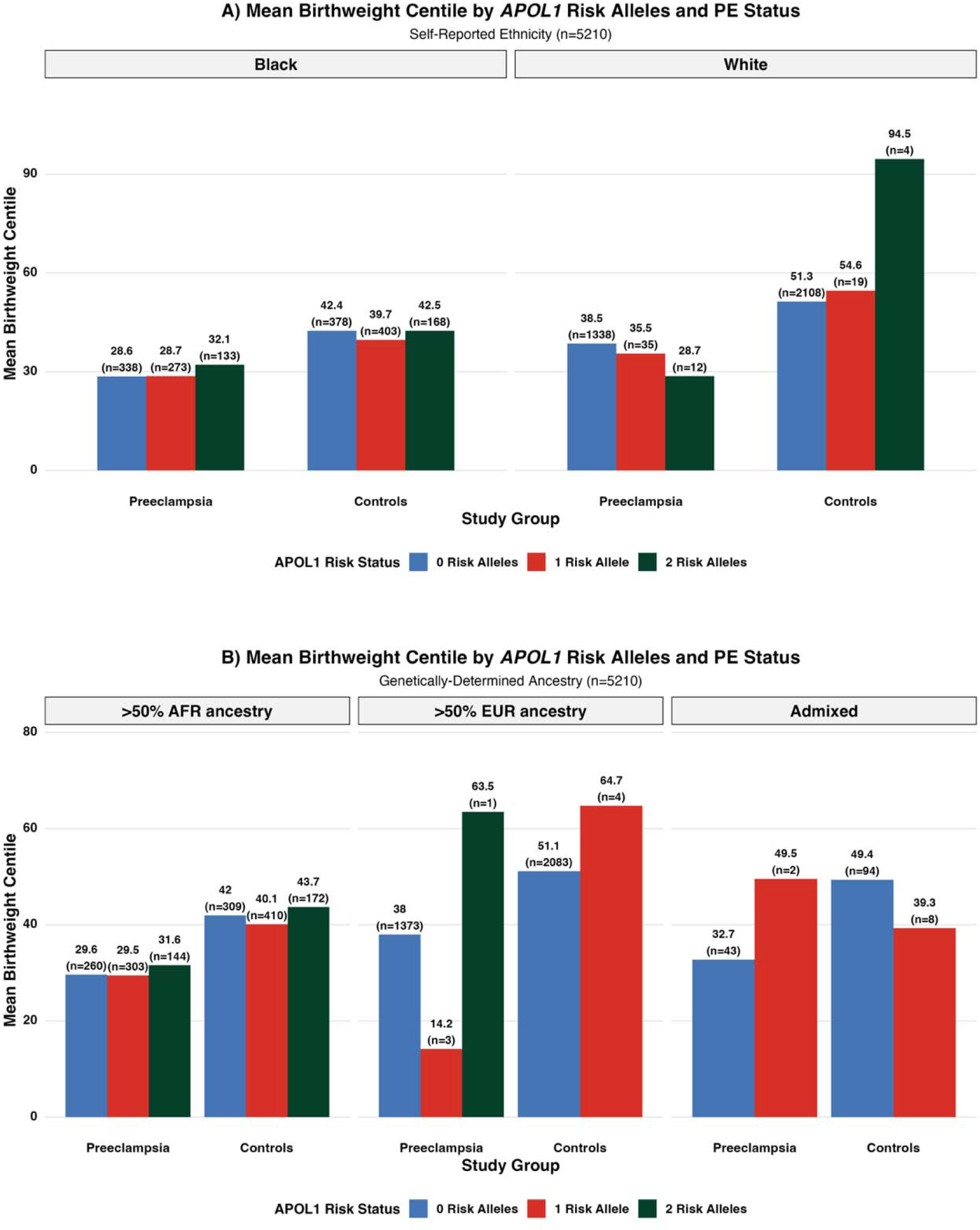
Distribution of Mean Birthweight Centile by *APOL1* Risk Alleles in Women with Preeclampsia and without Preeclampsia by Self-Reported Ethnicity (A) and by Genetically-Determined Ancestry. (B) Abbreviations: PE = preeclampsia, AFR = genetically-determined pan-African ancestry, EUR = genetically-determined European ancestry.

**Supplementary Figure S13.**
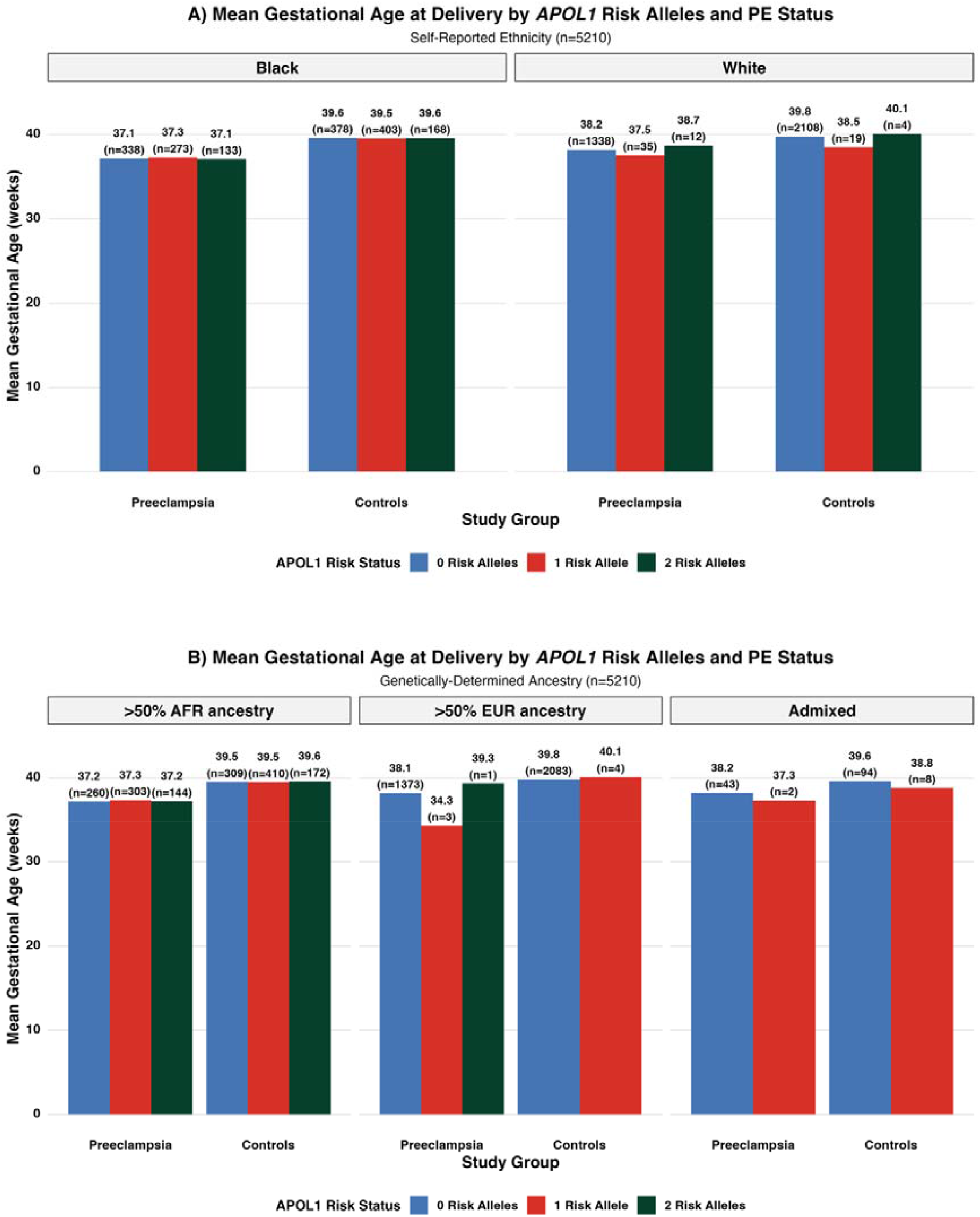
Distribution of Mean Gestational Age at Delivery by *APOL1* Risk Alleles in Women with Preeclampsia and without Preeclampsia by Self-Reported Ethnicity (A) and by Genetically-Determined Ancestry (B) Abbreviations: PE = preeclampsia, AFR = genetically-determined pan-African ancestry, EUR = genetically-determined European ancestry.

**Supplementary Figure S14.**
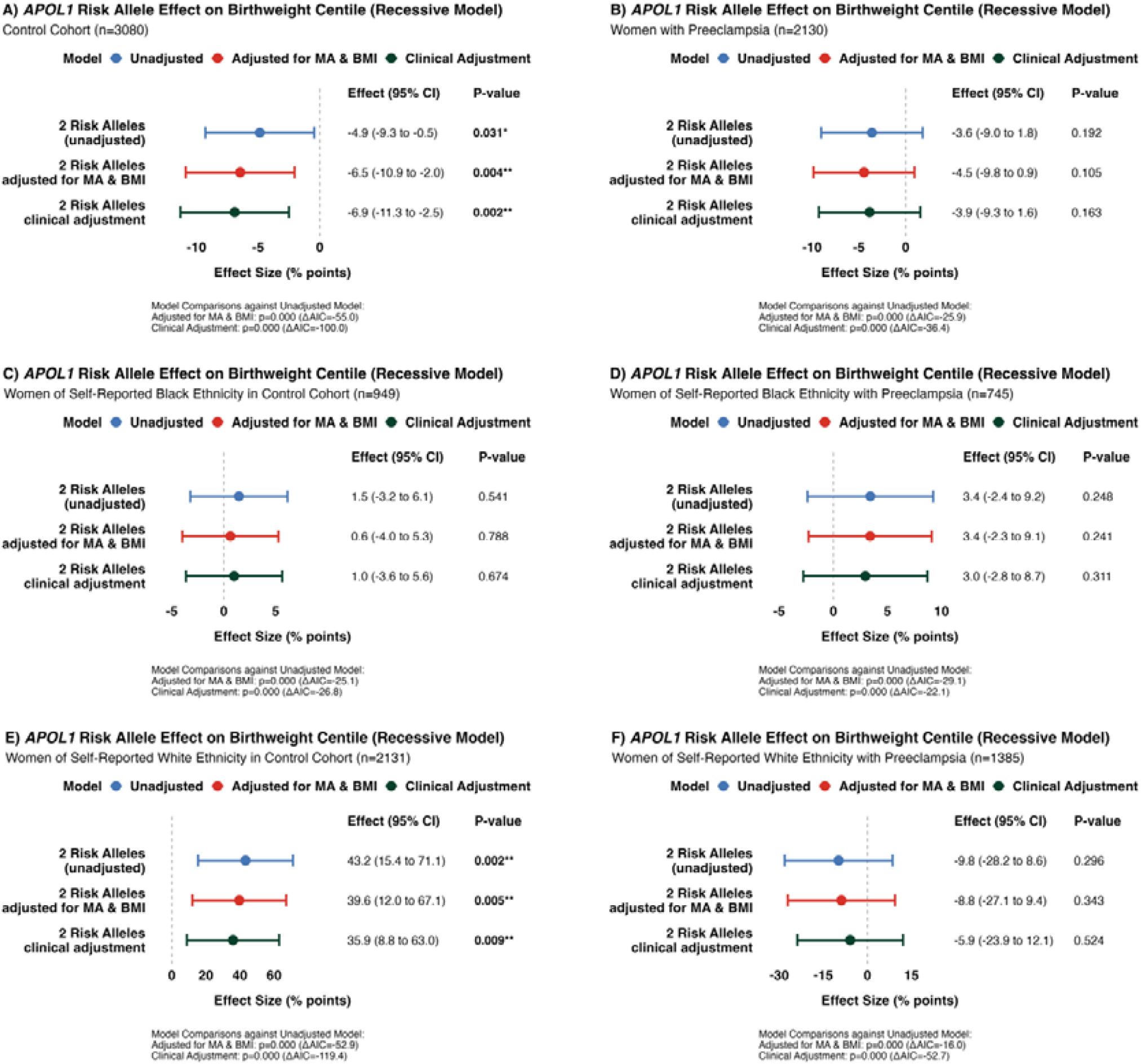
Effect of *APOL1* Risk Alleles on Birthweight Centile under the Recessive Model. Forest plots demonstrating effect size of presence of two *APOL1* risk alleles (G1G1, G1G2 or G2G2) compared to zero or one risk alleles (G0G0, G1G0 or G2G0) on birthweight in percentage points in the unadjusted model, model with adjustment for maternal age (MA) and body mass index (BMI) and model with clinical risk factor adjustment in the controls (A) and preeclampsia cases (B), in women with self-reported Black ethnic background in the controls(C) and with preeclampsia (D) and in women with self-reported White ethnic background in the controls (E) and with preeclampsia (F). Model fit and comparison was assessed using F-tests and the Akaike Information Criterion (AIC). CI = confidence interval.

**Supplementary Figure S15.**
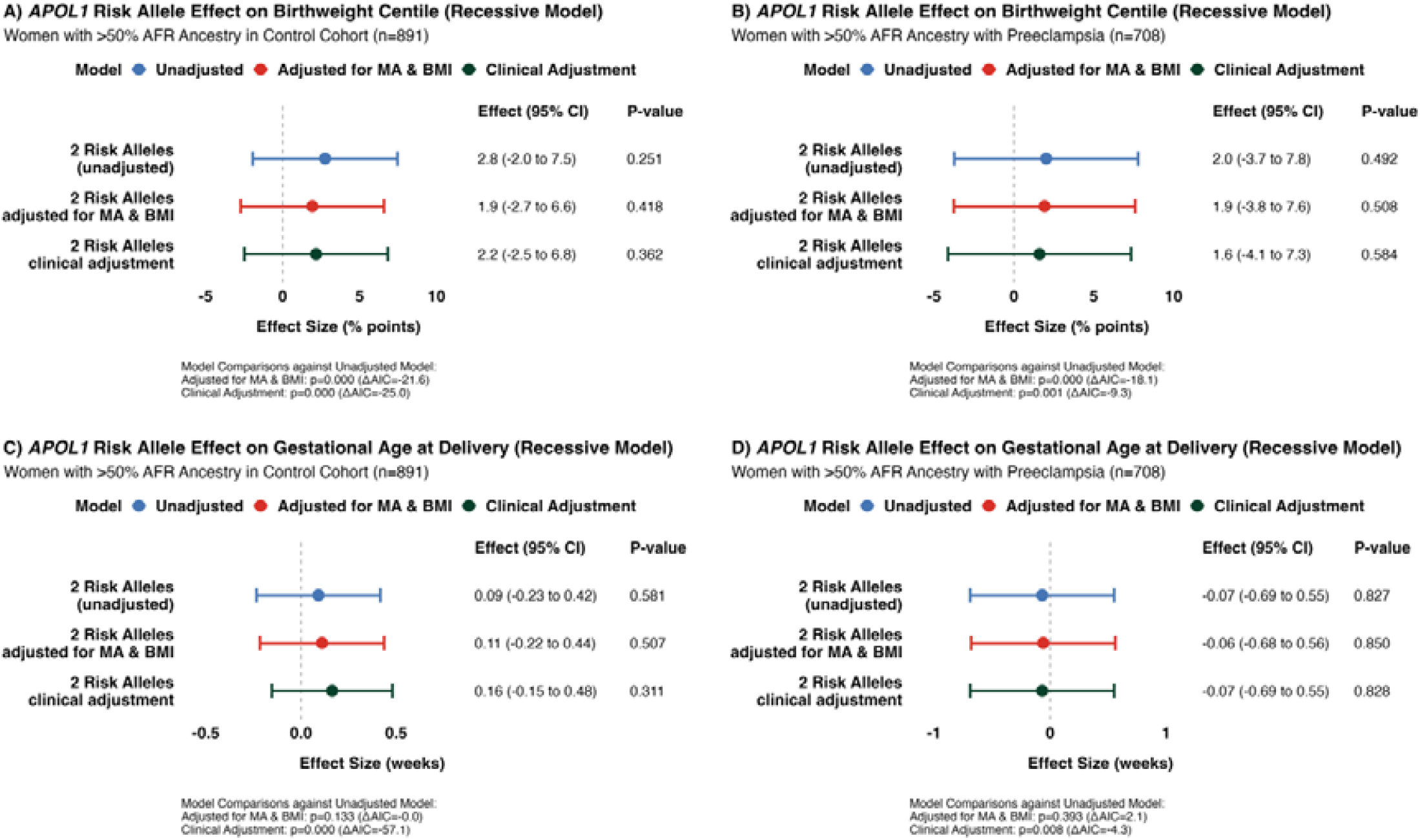
Effect of *APOL1* Risk Alleles on Birthweight Centile and Gestational Age at Delivery in Women with >50% pan-African Genetic Ancestry under the Recessive Model. Forest plots demonstrating effect size of presence of two *APOL1* risk alleles (G1G1, G1G2 or G2G2) compared to zero or one risk alleles (G0G0, G1G0 or G2G0) on birthweight in percentage points in the unadjusted model, model with adjustment for maternal age (MA) and body mass index (BMI) and model with clinical risk factor adjustment in women with >50% pan-African (AFR) genetic ancestry in the controls(A) and in the preeclampsia cases (B). Forest plots demonstrating effect size of presence of two *APOL1* risk alleles (G1G1, G1G2 or G2G2) compared to zero or one risk alleles (G0G0, G1G0 or G2G0) on gestational age at delivery in weeks in the unadjusted model, model with adjustment for maternal age (MA) and body mass index (BMI) and model with clinical risk factor adjustment in women with >50% pan-African (AFR) genetic ancestry in the controls (C) and in the preeclampsia cases (D). Model fit and comparison was assessed using F-tests and the Akaike Information Criterion (AIC). CI = confidence interval.

**Supplementary Figure S16.**
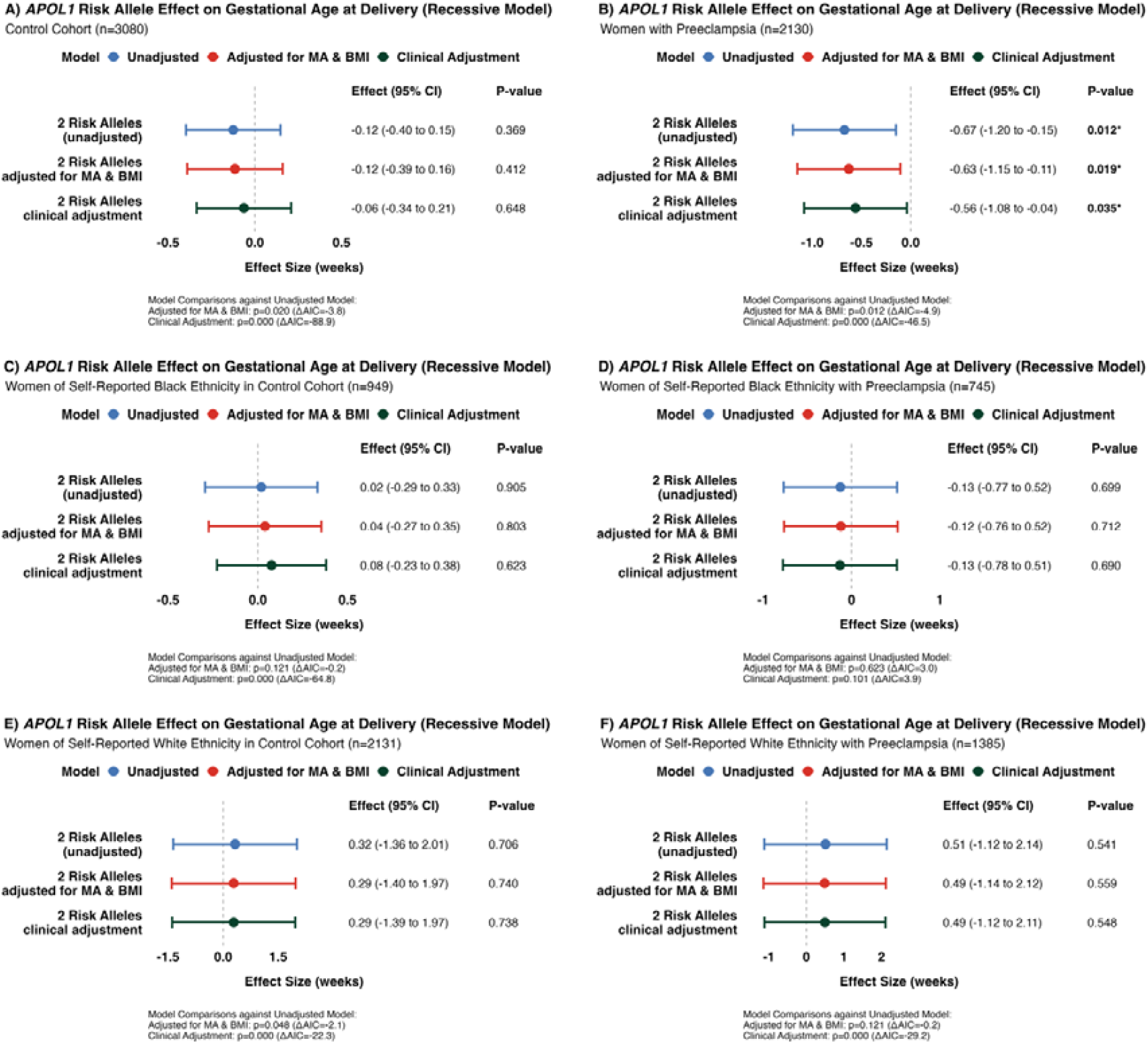
Effect of *APOL1* Risk Alleles on Gestational Age at Delivery under the Recessive Model. Forest plots demonstrating effect size of presence of two *APOL1* risk alleles (G1G1, G1G2 or G2G2) compared to zero or one risk alleles (G0G0, G1G0 or G2G0) on gestational age at delivery in weeks in the unadjusted model, model with adjustment for maternal age (MA) and body mass index (BMI) and model with clinical risk factor adjustment in the controls (A) and preeclampsia cases (B), in women with self-reported Black ethnic background in the controls(C) and with preeclampsia (D) and in women with self-reported White ethnic background in the controls (E) and with preeclampsia (F). Model fit and comparison was assessed using F-tests and the Akaike Information Criterion (AIC). CI = confidence interval.

## References

1. Poon, L.C., et al., The International Federation of Gynecology and Obstetrics (FIGO) initiative on pre-eclampsia: A pragmatic guide for first-trimester screening and prevention. Int J Gynaecol Obstet, 2019. 145 Suppl 1(Suppl 1): p. 1–33.

2. Maric-Bilkan, C., et al., Research Recommendations From the National Institutes of Health Workshop on Predicting, Preventing, and Treating Preeclampsia. Hypertension, 2019. 73(4): p. 757–766.

3. Force, U.P.S.T., Aspirin Use to Prevent Preeclampsia and Related Morbidity and Mortality: US Preventive Services Task Force Recommendation Statement. JAMA, 2021. 326(12): p. 1186–1191.

4. Fingar, K.R., et al., Delivery Hospitalizations Involving Preeclampsia and Eclampsia, 2005– 2014, in Healthcare Cost and Utilization Project (HCUP) Statistical Briefs. 2006, Agency for Healthcare Research and Quality (US): Rockville (MD).

5. Ghosh, A.P., et al., CHOP potentially co-operates with FOXO3a in neuronal cells to regulate PUMA and BIM expression in response to ER stress. PLoS One, 2012. 7(6): p. e39586.

6. Breathett, K., et al., Differences in Preeclampsia Rates Between African American and Caucasian Women: Trends from the National Hospital Discharge Survey. Journal of Women’s Health, 2014. 23(11): p. 886–893.

7. Ananth, C.V., K.M. Keyes, and R.J. Wapner, Pre-eclampsia rates in the United States, 1980-2010: age-period-cohort analysis. BMJ: British Medical Journal, 2013. 347: p. f6564.

8. Khalil, A., et al., Maternal racial origin and adverse pregnancy outcome: a cohort study. Ultrasound in Obstetrics & Gynecology, 2013. 41(3): p. 278–285.

9. Poon, L.C.Y., et al., Maternal risk factors for hypertensive disorders in pregnancy: a multivariate approach. Journal of Human Hypertension, 2010. 24(2): p. 104–110.

10. Force, U.P.S.T., Screening for Preeclampsia: US Preventive Services Task Force Recommendation Statement. JAMA, 2017. 317(16): p. 1661–1667.

11. Johnson, J.D. and J.M. Louis, Does race or ethnicity play a role in the origin, pathophysiology, and outcomes of preeclampsia? An expert review of the literature. American Journal of Obstetrics and Gynecology, 2022. 226(2, Supplement): p. S876–S885.

12. Ross, K.M., et al., Socioeconomic Status, Preeclampsia Risk and Gestational Length in Black and White Women. Journal of Racial and Ethnic Health Disparities, 2019. 6(6): p. 1182–1191.

13. Genovese, G., et al., Association of trypanolytic *APOL1* variants with kidney disease in African Americans. Science, 2010. 329(5993): p. 841–5.

14. Limou, S., et al., Sequencing rare and common <em>APOL1</em> coding variants to determine kidney disease risk. Kidney International, 2015. 88(4): p. 754–763.

15. Limou, S., et al., APOL1 kidney risk alleles: population genetics and disease associations. Adv Chronic Kidney Dis, 2014. 21(5): p. 426–33.

16. Kopp, J.B., et al., APOL1 Genetic Variants in Focal Segmental Glomerulosclerosis and HIV-Associated Nephropathy. Journal of the American Society of Nephrology, 2011. 22(11).

17. Tayo, B.O., et al., Genetic variation in *APOL1* and MYH9 genes is associated with chronic kidney disease among Nigerians. Int Urol Nephrol, 2013. 45(2): p. 485–94.

18. Ulasi, II, et al., High population frequencies of *APOL1* risk variants are associated with increased prevalence of non-diabetic chronic kidney disease in the Igbo people from south-eastern Nigeria. Nephron Clin Pract, 2013. 123(1-2): p. 123–8.

19. Tzur, S., et al., Missense mutations in the *APOL1* gene are highly associated with end stage kidney disease risk previously attributed to the MYH9 gene. Hum Genet, 2010. 128(3): p. 345–50.

20. Page, N.M., et al., The human apolipoprotein L gene cluster: identification, classification, and sites of distribution. Genomics, 2001. 74(1): p. 71–8.

21. Wen, Q., et al., Peptidomic Identification of Serum Peptides Diagnosing Preeclampsia. PLoS One, 2013. 8(6): p. e65571.

22. Bruggeman, L.A., et al., APOL1-G0 or APOL1-G2 Transgenic Models Develop Preeclampsia but Not Kidney Disease. J Am Soc Nephrol, 2016. 27(12): p. 3600–3610.

23. Sedor, J.R., L.A. Bruggeman, and J.F. O’Toole, APOL1 and Preeclampsia: Intriguing Links, Uncertain Causality, Troubling Implications. American Journal of Kidney Diseases, 2021. 77(6): p. 863–865.

24. Reidy, K.J., et al., Fetal-Not Maternal-APOL1 Genotype Associated with Risk for Preeclampsia in Those with African Ancestry. Am J Hum Genet, 2018. 103(3): p. 367–376.

25. Thakoordeen-Reddy, S., et al., Maternal variants within the apolipoprotein L1 gene are associated with preeclampsia in a South African cohort of African ancestry. Eur J Obstet Gynecol Reprod Biol, 2020. 246: p. 129–133.

26. Hong, X., et al., Joint Associations of Maternal-Fetal *APOL1* Genotypes and Maternal Country of Origin With Preeclampsia Risk. Am J Kidney Dis, 2021. 77(6): p. 879-888.e1.

27. American College of Obstetricians and Gynecologists: Gestational Hypertension and Preeclampsia. Obstetrics & Gynecology, 2020. 135:e237–e260.

28. Illumina. Infinium Global Screening Array-24 Kit. Accessed December 19, h.w.i.c.p.b.-t.m.-k.i.-g.-s.h.

29. Marees, A.T., et al., A tutorial on conducting genome-wide association studies: Quality control and statistical analysis. Int J Methods Psychiatr Res, 2018. 27(2): p. e1608.

30. Chang, C.C., et al., Second-generation PLINK: rising to the challenge of larger and richer datasets. GigaScience, 2015. 4(1).

31. Pritchard, J.K., M. Stephens, and P. Donnelly, Inference of population structure using multilocus genotype data. Genetics, 2000. 155(2): p. 945–59.

32. Alexander, D.H., J. Novembre, and K. Lange, Fast model-based estimation of ancestry in unrelated individuals. Genome Res, 2009. 19(9): p. 1655–64.

33. Conti-Ramsden, F., et al., Association of genetic ancestry with pre-eclampsia in multi-ethnic cohorts of pregnant women. Pregnancy Hypertension, 2024. 38: p. 101162.

34. Conti-Ramsden, F., et al., Genetically Estimated Ancestry and the Risk of Pre-Eclampsia. JACC: Advances, 2025. 4(12_Part_2): p. 102283.

35. Wright, D., A. Wright, and K.H. Nicolaides, The competing risk approach for prediction of preeclampsia. Am J Obstet Gynecol, 2020. 223(1): p. 12-23.e7.

36. Team, R.C., R: A language and environment for statistical computing. 2024, R Foundation for Statistical Computing: Vienna, Austria.

37. Magee, L.A., et al., The 2021 International Society for the Study of Hypertension in Pregnancy classification, diagnosis & management recommendations for international practice. Pregnancy Hypertens, 2022. 27: p. 148–169.

38. Nilsson, E., et al., The importance of genetic and environmental effects for pre-eclampsia and gestational hypertension: a family study. Bjog, 2004. 111(3): p. 200–6.

39. Cnattingius, S., et al., Maternal and fetal genetic factors account for most of familial aggregation of preeclampsia: a population-based Swedish cohort study. Am J Med Genet A, 2004. 130a(4): p. 365–71.

40. Thakoordeen, S., J. Moodley, and T. Naicker, Candidate Gene, Genome-Wide Association and Bioinformatic Studies in Pre-eclampsia: a Review. Current Hypertension Reports, 2018. 20(10).

41. Williams, P.J. and F. Broughton Pipkin, The genetics of pre-eclampsia and other hypertensive disorders of pregnancy. Best Practice & Research Clinical Obstetrics & Gynaecology, 2011. 25(4): p. 405–417.

42. Ardissino, M., et al., Polygenic Risk Scores for Preeclampsia Prediction Beyond Gold-Standard Clinical Models in Multiethnic Populations. J Am Heart Assoc, 2025. 14(24): p. e046211.

43. Honigberg, M.C., et al., Polygenic prediction of preeclampsia and gestational hypertension. Nat Med, 2023. 29(6): p. 1540–1549.

44. Conti-Ramsden, F., A. de Marvao, and L.C. Chappell, PREGNANCY DISORDERS AND MATERNAL CONSEQUENCES: Ethnic disparities in hypertensive disorders of pregnancy. Reproduction, 2025. 169(6): p. e250049.

45. Conti-Ramsden, F., et al., Does genetically-defined ancestry predict booking blood pressure in pregnant women with chronic hypertension? Pregnancy Hypertension, 2019. 17: p. S11.

46. Miller, A.K., et al., Association of preeclampsia with infant *APOL1* genotype in African Americans. BMC Med Genet, 2020. 21(1): p. 110.

47. Tzur, S., et al., APOL1 allelic variants are associated with lower age of dialysis initiation and thereby increased dialysis vintage in African and Hispanic Americans with non-diabetic end-stage kidney disease. Nephrol Dial Transplant, 2012. 27(4): p. 1498–505.

48. Kanji, Z., et al., Genetic variation in *APOL1* associates with younger age at hemodialysis initiation. J Am Soc Nephrol, 2011. 22(11): p. 2091–7.

49. Beckerman, P. and K. Susztak, APOL1: The Balance Imposed by Infection, Selection, and Kidney Disease. Trends Mol Med, 2018. 24(8): p. 682–695.

50. Huang, Q.Q., et al., Examining the role of common variants in rare neurodevelopmental conditions. Nature, 2024. 636(8042): p. 404–411.

51. Smail, C., et al., Complex trait associations in rare diseases and impacts on Mendelian variant interpretation. Nature Communications, 2024. 15(1): p. 8196.

52. Ma, L., J. Divers, and B.I. Freedman, Mechanisms of Injury in APOL1-associated Kidney Disease. Transplantation, 2019. 103(3): p. 487–492.

53. Kudose, S., et al., Kidney Biopsy Findings in Patients with COVID-19. J Am Soc Nephrol, 2020. 31(9): p. 1959–1968.

54. Wu, H., et al., AKI and Collapsing Glomerulopathy Associated with COVID-19 and APOL 1 High-Risk Genotype. J Am Soc Nephrol, 2020. 31(8): p. 1688–1695.

55. Daneshpajouhnejad, P., et al., The evolving story of apolipoprotein L1 nephropathy: the end of the beginning. Nat Rev Nephrol, 2022. 18(5): p. 307–320.

56. Yang, Y., et al., Interleukin-18 and interferon gamma levels in preeclampsia: a systematic review and meta-analysis. Am J Reprod Immunol, 2014. 72(5): p. 504–14.

57. Erez, O., et al., Preeclampsia and eclampsia: the conceptual evolution of a syndrome. Am J Obstet Gynecol, 2022. 226(2s): p. S786–s803.

